# Kynurenine pathway metabolites in cerebrospinal fluid and blood as potential biomarkers in Huntington’s disease

**DOI:** 10.1101/2020.08.06.20169524

**Authors:** Filipe B Rodrigues, Lauren M Byrne, Alexander J Lowe, Rosanna Tortelli, Mariette Heins, Gunnar Flik, Eileanoir B Johnson, Enrico De Vita, Rachael I Scahill, Flaviano Giorgini, Edward J Wild

## Abstract

**Background:** Converging lines of evidence from cell, yeast and animal models, and post-mortem human brain tissue studies, support the involvement of the kynurenine pathway (KP) in Huntington’s disease (HD) pathogenesis. Quantifying KP metabolites in HD biofluids is desirable, both to study pathobiology, and as a potential source of biomarkers to quantify pathway dysfunction and evaluate the biochemical impact of therapeutic interventions targeting its components.

**Methods:** In a prospective single-site controlled cohort study with standardised collection of CSF, blood, phenotypic and imaging data, we used high-performance liquid-chromatography to measure the levels of KP metabolites – tryptophan, kynurenine, kynurenic acid, 3-hydroxykynurenine, anthranilic acid and quinolinic acid – in CSF and plasma of 80 participants (20 healthy controls, 20 premanifest HD, and 40 manifest HD). We investigated short-term stability, intergroup differences, associations with clinical and imaging measures, and derived sample-size calculation for future studies.

**Findings:** Overall, KP metabolites in CSF and plasma were stable over 6 weeks, displayed no significant group differences and were not associated with clinical or imaging measures. Larger sample sizes would be needed to show differences in future studies.

**Interpretation:** We conclude that the studied metabolites are readily and reliably quantifiable in both biofluids in controls and HD gene expansion carriers. However, we found little evidence to support a substantial derangement of the KP in HD, at least to the extent that it is reflected by the levels of the metabolites in patient-derived biofluids.

**Fund:** This study was supported by the Medical Research Council UK and CHDI foundation.

**Research in Context Section:** *Evidence before this study:* The kynurenine pathway is a metabolic process needed for the degradation of tryptophan – an essential amino acid. Several by-products of this pathway have been implicated in the pathobiology of Huntington’s disease, a fatal neurodegenerative condition. Studying these metabolites could help better understand the biology of the condition and accelerate treatment development. In 2018, a systematic review concluded that only a small number of studies attempted to investigate the levels of these by-products in human biofluids, with the majority being limited by methodologic frailties and therefore requiring further study.

*Added value of this study:* We used a large prospective cohort consisting of Huntington’s disease mutation carriers and healthy controls to study the metabolic by-products of the kynurenine pathway. Matched cerebrospinal fluid and blood were collected using standardized protocol and analysed with high-performance liquid-chromatography. None of the studied metabolites showed associations with disease stage or with well-known clinical and imaging markers of the disease.

*Implication of all the available evidence:* Our results show that substantial alterations of the kynurenine pathway are not present in patients with Huntington’s disease compared to healthy controls, at least to the extent that is measurable in cerebrospinal fluid or blood. Whilst our results discourage the use of these metabolites as diagnostic and prognostic biomarkers, they do not reject the notion that regional- and tissue-specific alterations may exist, and that they may possess value as pharmacodynamic biomarkers in clinical trials targeting the kynurenine pathway.

## Introduction

Huntington’s disease (HD) is an invariably fatal neurodegenerative disease caused by CAG repeat expansions in the *HTT* gene. Inherited in an autosomal dominant manner, the polyglutamine expansion results in the ubiquitous expression of a mutant form of huntingtin protein (mHTT) (2), causing a diverse array of intracellular toxicities and derangement of downstream pathways. With no treatments shown to prevent, slow or reverse its progression (3), extensive dysfunction and neuronal death occurs. Although characterized by the degeneration of striatal medium spiny neurons (MSNs), widespread damage involving most brain regions is observed (4). In addition to its expression in glial cells and neurons, mHTT is expressed in the peripheral nervous system (5). The interplay amongst different cell types, both central and peripheral, and the dynamics of dysfunction versus compensation, are increasingly recognised as contributing to the complex pathogenesis of HD (6–8).

Early animal models used excitotoxins such as quinolinic acid (QUIN), injected into the striatum, to recapitulate specific cellular pathology and phenotypic features of HD (9, 10). Although influential, chemically-lesioned models have now largely been superseded by various transgenic animals (11). The observed selective vulnerability of striatal MSNs to such toxins led to the theory that excitotoxicity may be an inherent part of HD pathogenesis, a theory that remains of interest.

QUIN is produced in the central nervous system by microglial cells, as one endpoint of the kynurenine pathway (KP) of tryptophan (TRP) degradation (Figure 1 Overview of the kynurenine pathway. Adapted with authors and editors’ permission (1). The designations ‘neurotoxic’ and ‘neuroprotective’ are assigned on the basis of the balance of evidence and are acknowledged to be simplifications of complex properties.

Figure 1) (1), ultimately leading to the formation of the coenzyme nicotinamide adenine dinucleotide (NAD+). In addition to promoting excitotoxicity via activation of *N*-methyl-D-aspartate (NMDA) receptors, QUIN is a potent free radical generator (12). Several additional KP metabolites have been found to be neuroactive, including 3-hydroxykynurenine (3-HK) and kynurenic acid (KYNA). 3-HK is broadly neurotoxic via generation of free radicals (13–15), whereas KYNA displays neuroprotective properties via antagonism of excitatory amino acid receptors and scavenging of free radicals (16–19). The absolute levels of these metabolites, as well as the ratio of the neurotoxic compounds relative to KYNA, may be of value when considering progression of neurodegenerative disorders such as HD.

Furthermore, the activities of the key KP regulatory enzymes kynurenine-3-monooxygenase (KMO) – which synthesizes 3-HK leading to downstream formation of QUIN – and the kynurenine aminotransferases (KATs) which synthesize KYNA – are equally important (1).

**Figure 1.**
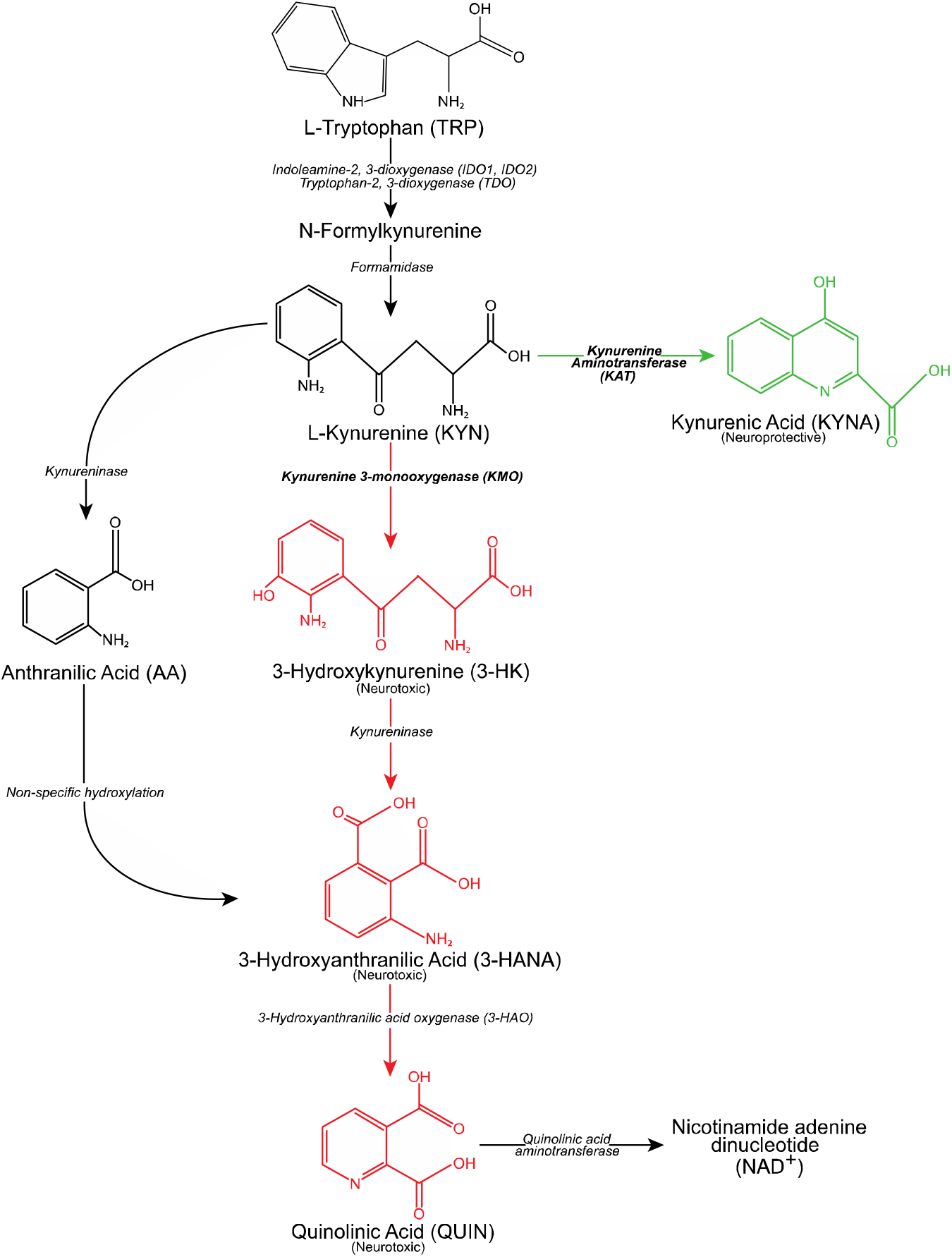
Overview of the kynurenine pathway. Adapted with authors and editors’ permission (1). The designations ‘neurotoxic’ and ‘neuroprotective’ are assigned on the basis of the balance of evidence and are acknowledged to be simplifications of complex properties.

Figure 2 Overview of the kynurenine pathway. Adapted with authors and editors’ permission (1). The designations ‘neurotoxic’ and ‘neuroprotective’ are assigned on the basis of the balance of evidence and are acknowledged to be simplifications of complex properties.

Converging lines of evidence support the involvement of the KP in HD pathogenesis. In post-mortem brain tissue obtained from HD patients, QUIN and 3-HK levels are increased and KYNA levels reduced (20–22). In the R6/2 mouse model, KMO activity has shown to be increased (23); however, recent work in a HD patient cohort failed to replicate these findings (Vonsattel neuropathology grades 1 through 4)(22). Furthermore, inhibition of KMO has demonstrated beneficial effects in multiple studies. In HD mouse models, peripheral KMO inhibition improves disease-relevant phenotypes (24, 25), while genetic inhibition is strongly protective in HD model yeast (26) and *Drosophila* (27–29). Consequently, pharmacological inhibition of KMO is considered a promising target for HD therapeutic development (1, 30).

Evidence of KP involvement in living human patients is limited. Blood and cerebrospinal fluid (CSF) are accessible biofluids that offer insights into comparative derangements in the periphery and CNS, and are sources of useful biomarkers in HD (31–33); however evidence on the KP in these fluids in HD is limited and conflicting. Early work examining CSF identified no difference in CSF QUIN levels in 10 HD patients versus 7 schizophrenic controls (34). This was further supported by Heyes *et al*. who observed no significant differences in CSF QUIN in 9 HD patients compared to 9 hospital patients using electron capture negative chemical ionization mass spectrometry and gas chromatography (35). In contrast with QUIN, levels of CSF kynurenine and KYNA were shown to be mildly reduced in 13 HD patients compared to 7 healthy controls (36).

Notably, all previous published human studies of the KP in HD CSF pre-date the discovery of the causative genetic mutation, used small sample numbers and were largely uncontrolled for important sources of variability like time of day, fasting status, CSF processing methods and source of control CSF(37).

Quantifying KP metabolites in HD CSF remains desirable, both to study pathobiology in human patients, and as a potential source of biomarkers to quantify pathway dysfunction and the biochemical impact of therapeutic interventions targeting its components. Therefore, we sought to combine modern analytical methods, with CSF and matched blood plasma, collected and processed under strictly standardised conditions, from a large (n = 80) prospective cohort of gene expansion carriers and matched controls (32, 38). Using high-performance liquid-chromatography (HPLC) with tandem mass spectroscopy (MS/MS) quantification methods, we measured levels of six KP metabolites (Figure 1 Overview of the kynurenine pathway. Adapted with authors and editors’ permission (1). The designations ‘neurotoxic’ and ‘neuroprotective’ are assigned on the basis of the balance of evidence and are acknowledged to be simplifications of complex properties.

Figure 1) – tryptophan (TRP), kynurenine (KYN), kynurenic acid (KYNA), 3-hydroxykynurenine (3-HK), anthranilic acid (AA) and quinolinic acid (QUIN) – in CSF and blood plasma, and applied a pre-defined statistical analysis plan to investigate the hypothesis that KMO activity is important to HD pathogenesis. As a pre-defined secondary analysis, we examined ratios of metabolites that may be informative of the activity of key enzymes in the pathway. Finally, we undertook an exploratory analysis of all metabolites in CSF and plasma in HD and controls.

## Methods

### Study design

The HD-CSF study was a prospective single-site controlled cohort study with standardised collection of CSF, blood and phenotypic data (online protocol: DOI: 10.5522/04/11828448.v1). Eighty participants were recruited (20 healthy controls, 20 premanifest HD [PreHD], and 40 manifest HD). All phenotypic assessment measures were predefined for HD-CSF based on metrics shown to have the largest effect sizes for predicting HD progression(39). Baseline assessments were conducted from February 2016 to February 2017(32). At baseline, 15 (19%) participants underwent an optional repeat sampling 4-8 weeks after baseline, permitting the assessment of within-subject short-term metabolite stability. MRI brain imaging, an optional component, was completed by 64 participants (80%).

### Ethical approval

This study was performed in accordance with the principles of the Declaration of Helsinki, and the International Conference on Harmonization Good Clinical Practice standards. Ethical approval was obtained from the London Camberwell St Giles Research Ethics Committee (15/LO/1917). Prior to undertaking study procedures, all participants gave informed consent which was obtained by clinical staff.

### Participants

Manifest HD participants were defined as adults having a Unified Huntington’s Disease Rating Scale (UHDRS) diagnostic confidence level (DCL) of 4 and *HTT* CAG repeat count ≥ 36. PreHD participants had CAG ≥ 40 and DCL < 4. Healthy controls were age- and gender-matched to gene expansion carriers, mostly spouses or gene-negative siblings of HD gene expansion carriers and with no neurological signs or symptoms. All were recruited from the National Hospital for Neurology & Neurosurgery Huntington’s Disease Service or the University College London Huntington’s Disease Centre research databases.

### Clinical assessments

Motor, cognitive and functional status were assessed using the UHDRS from the core Enroll-HD battery(40), including: the UHDRS Total Motor Score (TMS), Total Functional Capacity (TFC), Symbol Digit Modalities Test (SDMT), Stroop Word Reading (SWR), Stroop Color Naming (SCN) and Verbal Fluency – Categorical (VFC). These were performed at either a screening visit before sampling or an associated Enroll-HD visit (https://www.enroll-hd.org) within the 2 months prior to screening. We employed a calibrated iteration of the composite UHDRS (cUHDRS)(41, 42). Disease burden score (DBS) was calculated for each gene expansion carrier using the formula: [CAG – 35.5] × age(43). DBS estimates cumulative HD pathology as a function of CAG and the time exposed to the effects of the pathologic mutation, and has been shown to predict several features of disease progression including striatal pathology(39, 43).

### Biosample collection and processing

CSF and matched plasma were obtained as previous described (32). All collections were standardised for time of day after overnight fasting and processed within 30 minutes of collection using standardised equipment. Blood was collected within 10 minutes of CSF and processed to plasma. Biosamples were frozen and stored at −80°C until quantification.

### Kynurenine pathway metabolite quantification

KP metabolites were quantified by Charles River Laboratories (the Netherlands) using a high-performance liquid-chromatography (HPLC) with tandem mass spectroscopy (MS/MS) detection method (LC-MC) published previously (24). D_5_-TRP, D_4_-KYN, D_5_-KYNA, ^13^C_6_-3-HK, D_4_-AA and a D_3_-QUIN were used as internal standards. An aliquot of a solution containing the internal standard was mixed with an aliquot of each experimental sample to generate an LC-MS sample. An aliquot of each LC-MS sample was injected into the HPLC system by an automated sample injector (SIL20-AD, Shimadzu, Japan). Chromatographic separation was performed using a reversed phase analytical column, configured as per *Table 1*, with elution performed using a linear gradient. MS analyses were performed using an API 4000 MS/MS system consisting of an API 4000 MS/MS detector and a Turbo Ion Spray interface (Applied Biosystems, USA). The acquisitions on API 4000 were performed in positive ionization mode, with optimized settings for the analytes. The instrument was operated in multiple-reaction-monitoring (MRM) mode. Data were calibrated and quantified using the Analyst™ data system (Applied Biosystems, USA).

Assays were performed blinded to clinical data and in a continuous run with the same quality control samples (QCs) to confirm performance over time. Each sample was measured once. The acceptance criteria for CSF assays was +/-25% accuracy for the low limits of quantification (LoQ) calibrator and QC-low, and +/-20% accuracy for all other calibrators and QC-mid and -high. For plasma assays, acceptance criteria were 5% wider. Individual runs were accepted when > 66% of QC’s were within criteria as described above (> 50% on individual level). The LoQ, frequency of samples below LoQ, and frequency of rejected samples due to quality criteria are shown in *Table 1*.

**Table 1.** Chromatographic separation parameters for each metabolite and assay performance. Note that 2 manifest HD participants are missing plasma. 3-HK, 3-hydroxykynurenine; AA, anthranilic acid; ACN; acetonitrile; C, cerebrospinal fluid; FA, formic acid; H_2_O, ultra-purified water; KYN, kynurenine; KYNA, kynurenic acid; P, plasma; QUIN, quinolinic acid; TFA, trifluoroacetic acid; TRP, tryptophan.

| Metabolite | Column size (mm) | Particle size (µm) | Temperature (°C) | Mobile phase A | Mobile phase B | Elution flow rate (ml/min) | Low limit of quantification (LoQ) | n (%) below LoQ | n (%) quality rejected |
| --- | --- | --- | --- | --- | --- | --- | --- | --- | --- |
| TRP | 100 x 3 | 2.5 | 5 | H<sub>2</sub>O + 0.1% FA | ACN + 0.1% FA | 0.3 | C: 500 nM<br>P: 500 nM | C: 0/95 (0%)<br>P: 0/93 (0%) | C: 0/95 (0%)<br>P: 0/93 (0%) |
| KYN | 150 x 2.1 | 3 | 25 | H<sub>2</sub>O + 0.1% FA | ACN + 0.1% FA | 0.2 | C: 2.5 nM<br>P: 0.5 µM | C: 0/95 (0%)<br>P: 0/93 (0%) | C: 0/95 (0%)<br>P: 0/93 (0%) |
| KYNA | 150 x 2.1 | 3 | 25 | H<sub>2</sub>O + 0.1% FA | ACN + 0.1% FA | 0.2 | C: 0.25 nM<br>P: 15.0 nM | C: 0/95 (0%)<br>P: 4/93 (4%) | C: 0/95 (0%)<br>P: 4/93 (4%) |
| 3-HK | 150 x 2.1 | 3 | 25 | H<sub>2</sub>O + 0.1% FA | ACN + 0.1% FA | 0.2 | C: 0.25 nM<br>P: 5.0 nM | C: 0/95 (0%)<br>P: 1/93 (1%) | C: 0/95 (0%)<br>P: 0/93 (0%) |
| AA | 150 x 2.1 | 3 | 25 | H<sub>2</sub>O + 0.1% FA | ACN + 0.1% FA | 0.2 | C: 1.25 nM<br>P: 5.0 nM | C: 0/95 (0%)<br>P: 7/93 (8%) | C: 0/95 (0%)<br>P: 0/93 (0%) |
| QUIN | 100 x 3 | 2.5 | 35 | H<sub>2</sub>O + 0.2% TFA | ACN + 0.2% TFA | 0.3 | C: 1.0 nM<br>P: 100 nM | C: 0/95 (0%)<br>P: 1/93 (1%) | C: 0/95 (0%)<br>P: 0/93 (0%) |

### MRI Acquisition

T1-weighted MRI data were acquired on a 3T Siemens Prisma scanner using a protocol optimized for this study. Images were acquired using a 3D magnetization-prepared 180 degrees radio-frequency pulses and rapid gradient-echo (MPRAGE) sequence with a repetition time (TR) =2000 ms and echo time (TE)=2.05 ms. The protocol had an inversion time of 850 ms, flip angle of 8 degrees, matrix size 256 × 240 mm. 256 coronal partitions were collected to cover the entire brain with a slice thickness of 1·0 mm. Parallel imaging acceleration (GeneRalized Autocalibrating Partial Parallel Acquisition [GRAPPA], acceleration factor [R]=2) was used and 3D distortion correction was applied to all images.

### MRI Processing

All T1-weighted scans passed visual quality control check for the presence of significant motion or other artefacts before processing. Bias correction was performed using the N3 procedure (44). A semiautomated segmentation procedure via Medical Image Display Analysis Software (MIDAS) was used to generate volumetric regions of the whole brain and total intracranial volume (TIV), as previously described (45–47). SPM12 “Segment” (MATLAB version 2012b) was used to measure the volume of the grey and white matter (48). Multi-Atlas Propagation with EM Refinement (MALP-EM) was used to quantify caudate volume (49). MALP-EM is an automated tool used to segment MRI scans into regional volumes and has previously been validated for use in HD cohorts (50). Default settings were used for both SPM12 segmentations and MALP-EM caudate regions. No scans failed processing after visual quality control of segmentations by experienced raters to ensure accurate delineation of the regions. Baseline MRI volumes were presented adjusted for TIV. All MRI analyses used brain volumes as percentage of TIV.

### Statistical analyses

Statistical analysis was performed with Stata MP 16 software (StataCorp, USA). To minimise type 1 errors (i.e. rejection of true null hypotheses) resulting from multiplicity, we pre-defined CSF levels of KYNA, 3-HK and QUIN as primary outcomes, hypothesising that quantifiable differences would be apparent if the KP is dysregulated in HD. The ratios of CSF 3-HK:KYNA, KYNA:KYN and 3-HK:KYN were secondary outcomes, hypothesised to reflect the overall balance of the key neuroprotective and neurotoxic metabolites, the activity of the KATs, and the activity of KMO respectively (Figure 1 Overview of the kynurenine pathway. Adapted with authors and editors’ permission (1). The designations ‘neurotoxic’ and ‘neuroprotective’ are assigned on the basis of the balance of evidence and are acknowledged to be simplifications of complex properties.

Figure 1). Analyses of all other CSF and plasma metabolites were exploratory. Measurements below the LoD for the metabolites of interest were assumed as missing – no imputation was used. Analyte distributions were visually assessed and if necessary, arithmetical transformations were applied to meet model assumptions (Figure S1–3). Continuous variables were reported as mean ± standard deviations (SD); and categorical variables as absolute (n) and relative frequencies (%). Associations with age (healthy controls only), gender, CAG repeat count (gene expansion carriers only) and haemoglobin (in case of CSF metabolites only) and time in freezer storage were visually assessed for all outcomes (Figure S4–6). Due to its known effects on HD, all models included age as covariate.

As a planned second-level analysis, to assess associations with measures beyond the known combined effect of age and *HTT* CAG repeat count, models including gene expansion carriers only were run with age and CAG repeat count as covariates. To investigate intergroup differences in outcomes of interest, we applied generalised linear regression models estimated via ordinary least squares. Models were fitted with the variable of interest as the dependent variable, and first with age and group membership, and then with age and CAG and group membership as independent variables. We report results for the omnibus group membership main effect test and relevant contrasts (i.e. healthy controls vs PreHD and PreHD vs manifest HD). Other comparisons (i.e. healthy controls vs manifest HD) were not undertaken as we treat HD as a biological continuum. To investigate alterations unique to later stages of the disease we used clinical measures of disease progression (cUHDRS) and/or cumulative mHTT toxicity (DBS). Tests were not adjusted for multiple comparisons but the statistical approach was designed to minimise multiplicity within each question of independent interest.

We used Pearson’s partial correlations adjusted for age, and for age and CAG to study associations between outcomes of interest and DBS, clinical and imaging measures in gene expansion carriers. Pearson’s correlations were used to study associations between metabolites in CSF and plasma. Bias-corrected and accelerated bootstrapped 95% confidence intervals (95% CI) were calculated for correlation coefficients. We calculated interclass-correlation coefficients (ICC) and 95% CI using two-way mixed-effects models to investigate within-subject short-term stability. Sample size calculations for two 2-group comparisons (i.e. healthy control vs PreHD and PreHD vs manifest HD) using 2-sided Student t-tests were performed to inform future studies using the detected effect sizes adjusted for age in primary and secondary outcomes, and assuming a 5% type 1 error and 20% type 2 error (80% power). No adjustments were made for multiplicity.

## Results

### Demographics

Eighty participants were recruited – all contributed with CSF and 2 (3%) manifest HD were missing plasma. Full cohort characteristics are presented in Table S1. Disease groups were well-matched for gender and differed as expected in clinical, cognitive and imaging measures. Age differed significantly between groups due to the control group (50·68 years ± 11·0) being matched to all gene expansion carriers, and manifest HD (56·02 years ± 9·36) being more advanced in their disease course than preHD (42·38 years ± 11·04), as previously reported(32, 38).

### Primary outcomes

All three primary outcomes – CSF KYNA, 3-HK and QUIN – were associated with age in healthy controls, but not with gender, CAG repeat count (gene expansion carriers only), CSF haemoglobin or time in freezer storage (Figure S4). Age- and age and CAG-adjusted CSF levels of KYNA, 3-HK and QUIN showed no significant difference between groups *(Figure 3; Table 2)*.

**Figure 3.**
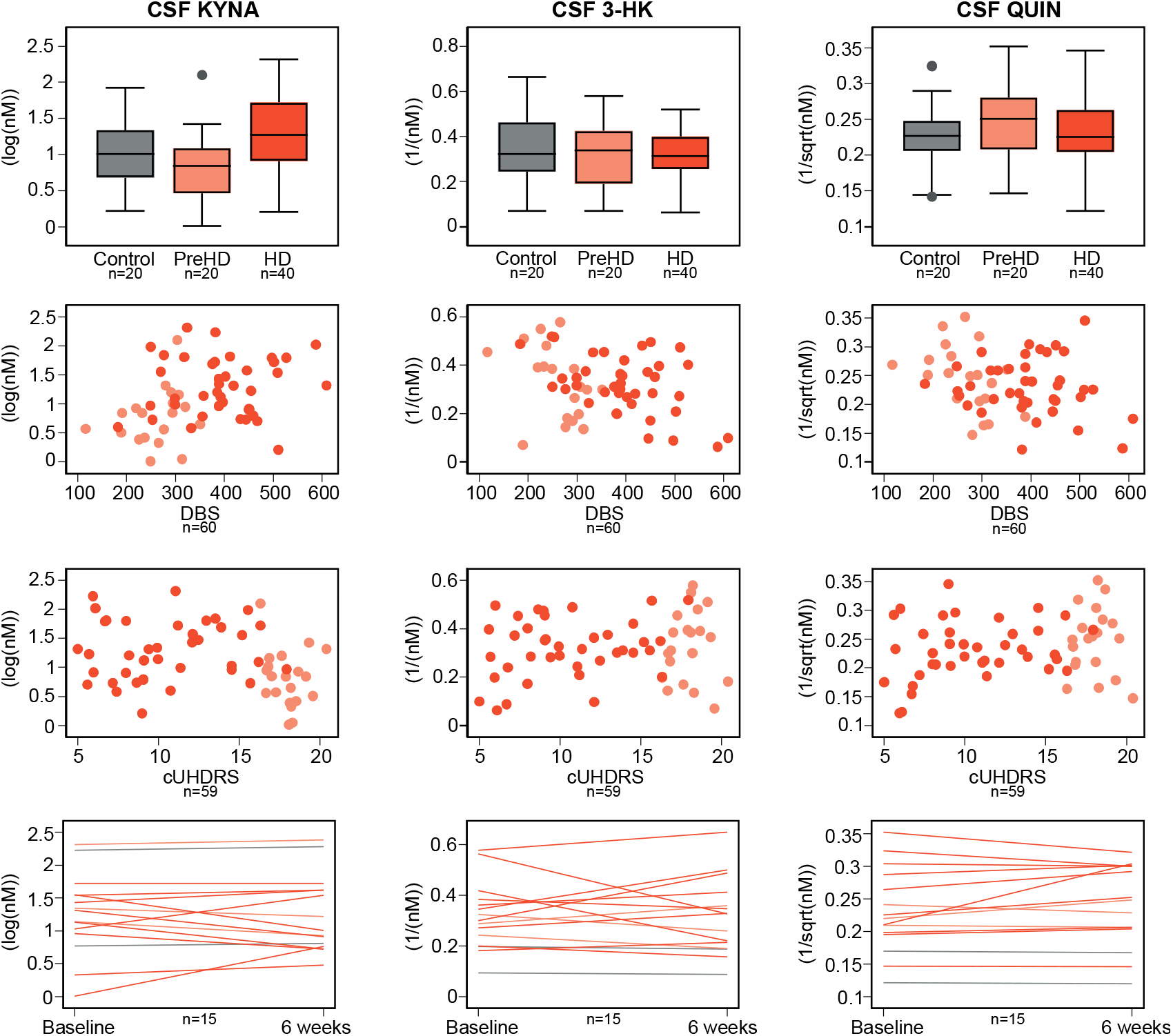
Intergroup differences (top row), associations with Disease Burden Score (DBS, second row) and composite Unified Huntington’s Disease Rating Scale (cUHDRS, third row), and within-subject short-term stability (bottom row) for primary outcomes: cerebrospinal fluid (CSF) kynurenic acid (KYNA), CSF 3-hydroxykynurenine (3-HK) and CSF quinolinic acid (QUIN). Associations were not found in any of the analyses. Grey represents healthy controls, light orange preHD and dark orange HD.

**Table 2.**
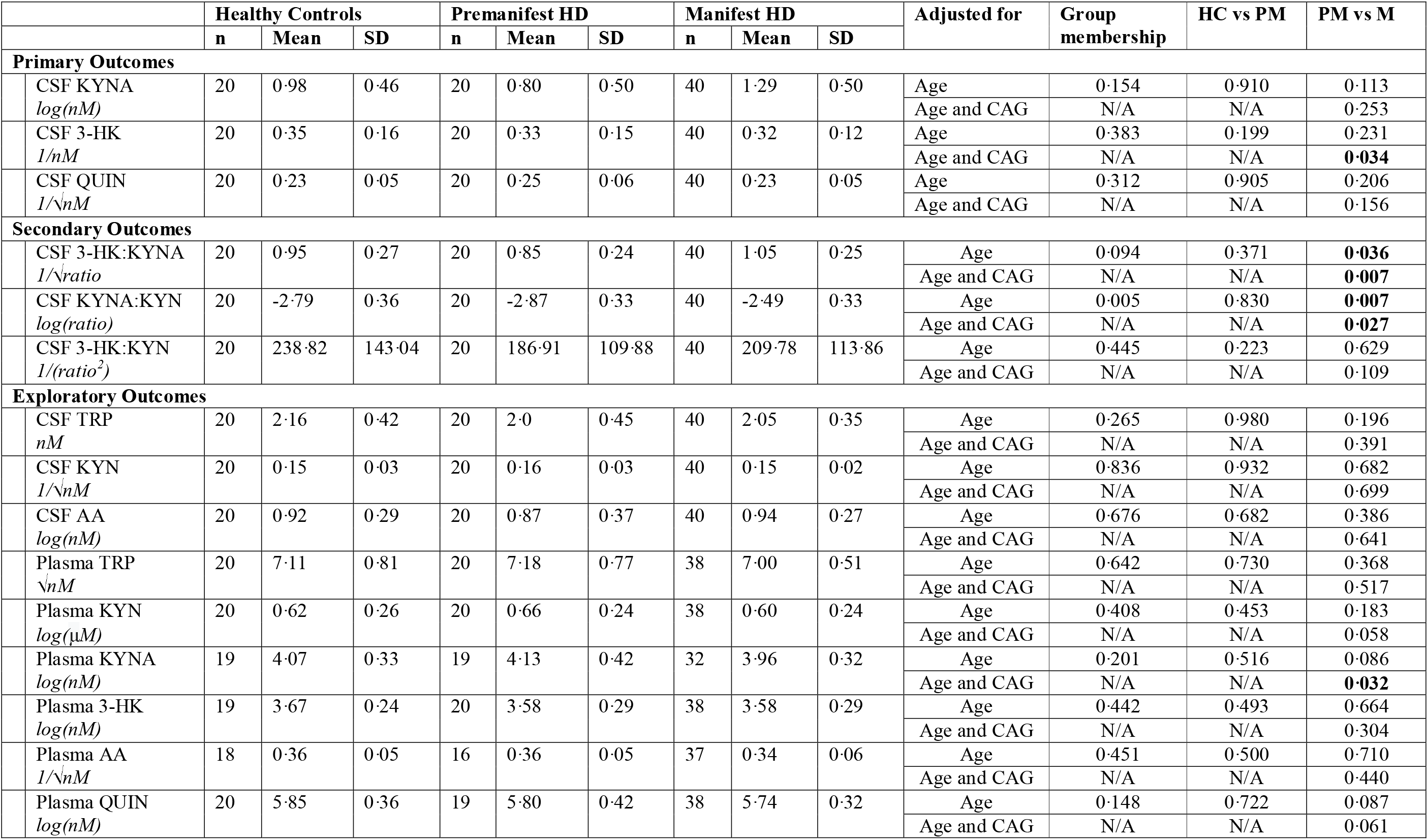
Intergroup comparisons. Note that for outcomes where a transformation including a reciprocal (i. e. 1/x), an increase in the raw measurement corresponds to a decreased in the transformed value and vice-versa. 3-HK, 3-hydroxykynurenine; AA, Anthranilic acid; CSF, cerebrospinal fluid; HC, healthy controls; KYN, kynurenine; KYNA, kynurenic acid; M, manifest HD; N/A, not applicable; PM, premanifest HD; QUIN, quinolinic acid; SD, standard deviation; TRP, tryptophan.

|  |  | Healthy Controls |  |  | Premanifest HD |  |  | Manifest HD |  |  | Adjusted for | Group membership | HC vs PM | PM vs M |
| --- | --- | --- | --- | --- | --- | --- | --- | --- | --- | --- | --- | --- | --- | --- |
|  |  | n | Mean | SD | n | Mean | SD | n | Mean | SD |  |  |  |  |
| Primary Outcomes |  |  |  |  |  |  |  |  |  |  |  |  |  |  |
|  | CSF KYNA<br>*log(nM)* | 20 | 0.98 | 0.46 | 20 | 0.80 | 0.50 | 40 | 1.29 | 0.50 | Age | 0.154 | 0.910 | 0.113 |
|  |  |  |  |  |  |  |  |  |  |  | Age and CAG | N/A | N/A | 0.253 |
|  | CSF 3-HK<br>*1/nM* | 20 | 0.35 | 0.16 | 20 | 0.33 | 0.15 | 40 | 0.32 | 0.12 | Age | 0.383 | 0.199 | 0.231 |
|  |  |  |  |  |  |  |  |  |  |  | Age and CAG | N/A | N/A | **0.034** |
|  | CSF QUIN<br>*1/√nM* | 20 | 0.23 | 0.05 | 20 | 0.25 | 0.06 | 40 | 0.23 | 0.05 | Age | 0.312 | 0.905 | 0.206 |
|  |  |  |  |  |  |  |  |  |  |  | Age and CAG | N/A | N/A | 0.156 |
| Secondary Outcomes |  |  |  |  |  |  |  |  |  |  |  |  |  |  |
|  | CSF 3-HK:KYNA<br>*1/√ratio* | 20 | 0.95 | 0.27 | 20 | 0.85 | 0.24 | 40 | 1.05 | 0.25 | Age | 0.094 | 0.371 | **0.036** |
|  |  |  |  |  |  |  |  |  |  |  | Age and CAG | N/A | N/A | **0.007** |
|  | CSF KYNA:KYN<br>*log(ratio)* | 20 | -2.79 | 0.36 | 20 | -2.87 | 0.33 | 40 | -2.49 | 0.33 | Age | 0.005 | 0.830 | **0.007** |
|  |  |  |  |  |  |  |  |  |  |  | Age and CAG | N/A | N/A | **0.027** |
|  | CSF 3-HK:KYN<br>*1/(ratio<sup>2</sup>)* | 20 | 238.82 | 143.04 | 20 | 186.91 | 109.88 | 40 | 209.78 | 113.86 | Age | 0.445 | 0.223 | 0.629 |
|  |  |  |  |  |  |  |  |  |  |  | Age and CAG | N/A | N/A | 0.109 |
| Exploratory Outcomes |  |  |  |  |  |  |  |  |  |  |  |  |  |  |
|  | CSF TRP<br>*nM* | 20 | 2.16 | 0.42 | 20 | 2.0 | 0.45 | 40 | 2.05 | 0.35 | Age | 0.265 | 0.980 | 0.196 |
|  |  |  |  |  |  |  |  |  |  |  | Age and CAG | N/A | N/A | 0.391 |
|  | CSF KYN<br>*1/√nM* | 20 | 0.15 | 0.03 | 20 | 0.16 | 0.03 | 40 | 0.15 | 0.02 | Age | 0.836 | 0.932 | 0.682 |
|  |  |  |  |  |  |  |  |  |  |  | Age and CAG | N/A | N/A | 0.699 |
|  | CSF AA<br>*log(nM)* | 20 | 0.92 | 0.29 | 20 | 0.87 | 0.37 | 40 | 0.94 | 0.27 | Age | 0.676 | 0.682 | 0.386 |
|  |  |  |  |  |  |  |  |  |  |  | Age and CAG | N/A | N/A | 0.641 |
|  | Plasma TRP<br>*√nM* | 20 | 7.11 | 0.81 | 20 | 7.18 | 0.77 | 38 | 7.00 | 0.51 | Age | 0.642 | 0.730 | 0.368 |
|  |  |  |  |  |  |  |  |  |  |  | Age and CAG | N/A | N/A | 0.517 |
|  | Plasma KYN<br>*log(μM)* | 20 | 0.62 | 0.26 | 20 | 0.66 | 0.24 | 38 | 0.60 | 0.24 | Age | 0.408 | 0.453 | 0.183 |
|  |  |  |  |  |  |  |  |  |  |  | Age and CAG | N/A | N/A | 0.058 |
|  | Plasma KYNA<br>*log(nM)* | 19 | 4.07 | 0.33 | 19 | 4.13 | 0.42 | 32 | 3.96 | 0.32 | Age | 0.201 | 0.516 | 0.086 |
|  |  |  |  |  |  |  |  |  |  |  | Age and CAG | N/A | N/A | **0.032** |
|  | Plasma 3-HK<br>*log(nM)* | 19 | 3.67 | 0.24 | 20 | 3.58 | 0.29 | 38 | 3.58 | 0.29 | Age | 0.442 | 0.493 | 0.664 |
|  |  |  |  |  |  |  |  |  |  |  | Age and CAG | N/A | N/A | 0.304 |
|  | Plasma AA<br>*1/√nM* | 18 | 0.36 | 0.05 | 16 | 0.36 | 0.05 | 37 | 0.34 | 0.06 | Age | 0.451 | 0.500 | 0.710 |
|  |  |  |  |  |  |  |  |  |  |  | Age and CAG | N/A | N/A | 0.440 |
|  | Plasma QUIN<br>*log(nM)* | 20 | 5.85 | 0.36 | 19 | 5.80 | 0.42 | 38 | 5.74 | 0.32 | Age | 0.148 | 0.722 | 0.087 |
|  |  |  |  |  |  |  |  |  |  |  | Age and CAG | N/A | N/A | 0.061 |

Associations between age-adjusted CSF levels of KYNA, 3-HK or QUIN and DBS, clinical and imaging measures were negligible to weak (*Figure 4*, Table S2 and Figure S8).

**Figure 4.**
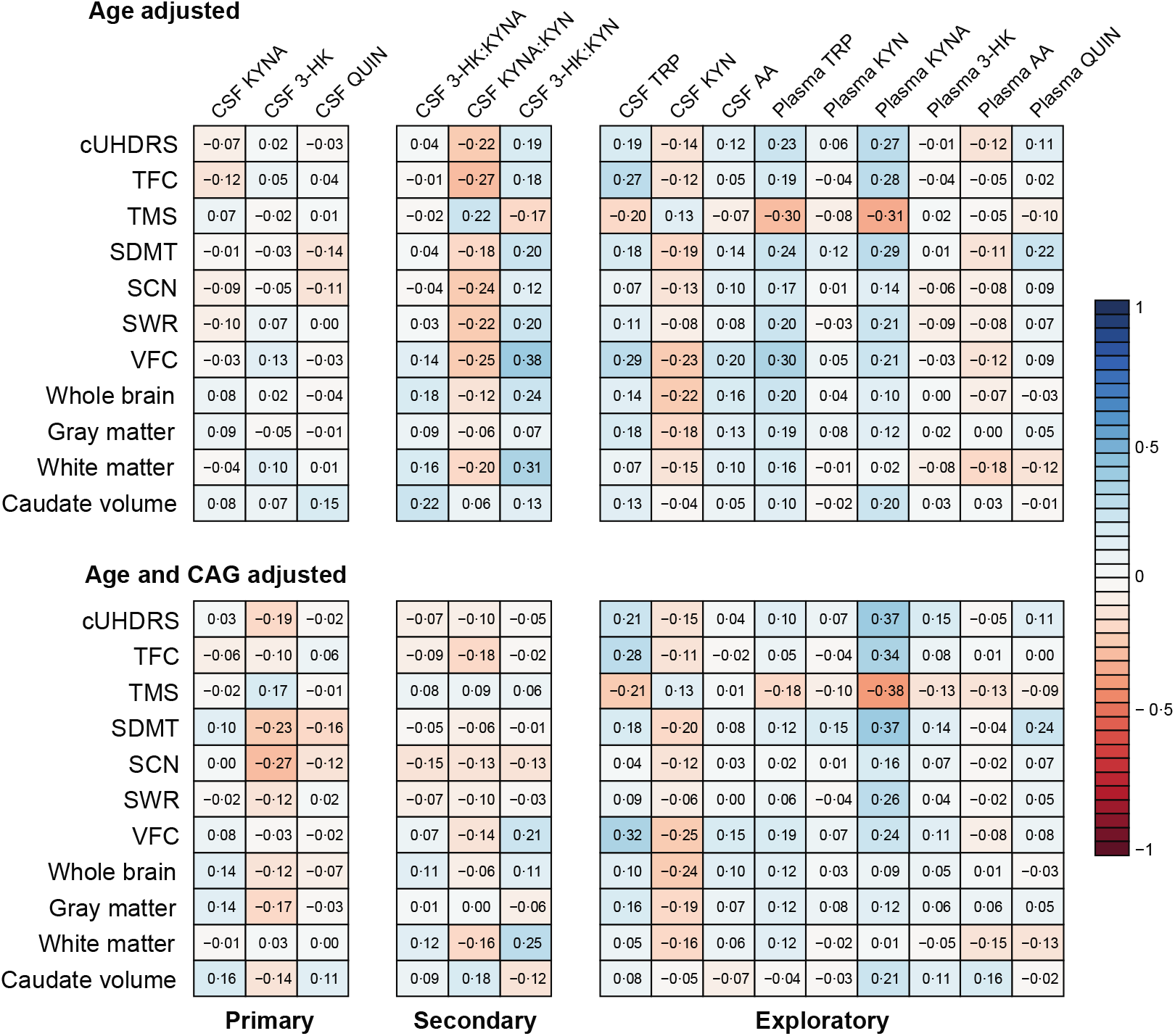
Associations between outcomes and clinical and imaging measures for primary, secondary and exploratory outcomes in gene expansion carriers. Numbers are Pearson s partial correlations coefficients adjusted for age (top row), and for age and CAG (bottom row; see Table S2 for 95% CI and Figure S8 for scatter plots). 3-HK, 3-hydroxykynurenine; AA, Anthranilic acid; CSF, cerebrospinal fluid; cUHDRS, composite Unified Huntington’s Disease Rating Scale; KYN, kynurenine; KYNA, kynurenic acid; QUIN, quinolinic acid; SCN, Stroop Color Naming’ SDMT, Symbol Digit Modalities Test; SWR, Stroop Word Reading’ TFC, UHDRS Total Functional Capacity; TMS, UHDRS Total Motor Score; TRP, tryptophan; VFC, Verbal Fluency – Categorical.

Within-subject short-term stability was good for CSF KYNA (ICC 0·84, 95% CI 0·58 to 0·94) and QUIN (ICC 0·90, 95% CI 0·72 to 0·96), and moderate for 3-HK (ICC 0·68, 95% CI 0·27 to 0·88, *Figure 3* and Table S3).

Power calculations showed that the number of participants per arm needed to give 80% power to detect a difference at the alpha level of 0·05 in age-adjusted CSF levels between healthy controls and preHD would be 48,921 for KYNA, 376 for 3-HK and 43,665 for QUIN. For the comparison between preHD and HD the number would be 368 for KYNA, 648 for 3-HK and 579 for QUIN.

### Secondary outcomes

None of the three pre-defined ratios of interest – CSF 3-HK:KYNA, KYNA:KYN and 3-HK:KYN – were associated with age, gender, CAG repeat count, CSF haemoglobin or time in the freezer (Figure S5). Age- and age and CAG-adjusted CSF levels of 3-HK:KYNA, KYNA:KYN and 3-HK:KYN showed no difference between healthy controls and preHD *(Figure* 5; *Table 2)*. There were differences in ratios of 3-HK:KYNA and KYNA:KYN between preHD and manifest HD (age-adjusted p-values = 0·036 and 0·007, respectively; age- and CAG-adjusted p-values = 0·007 and 0·027, respectively). No differences were found for 3-HK:KYN.

**Figure 5.**
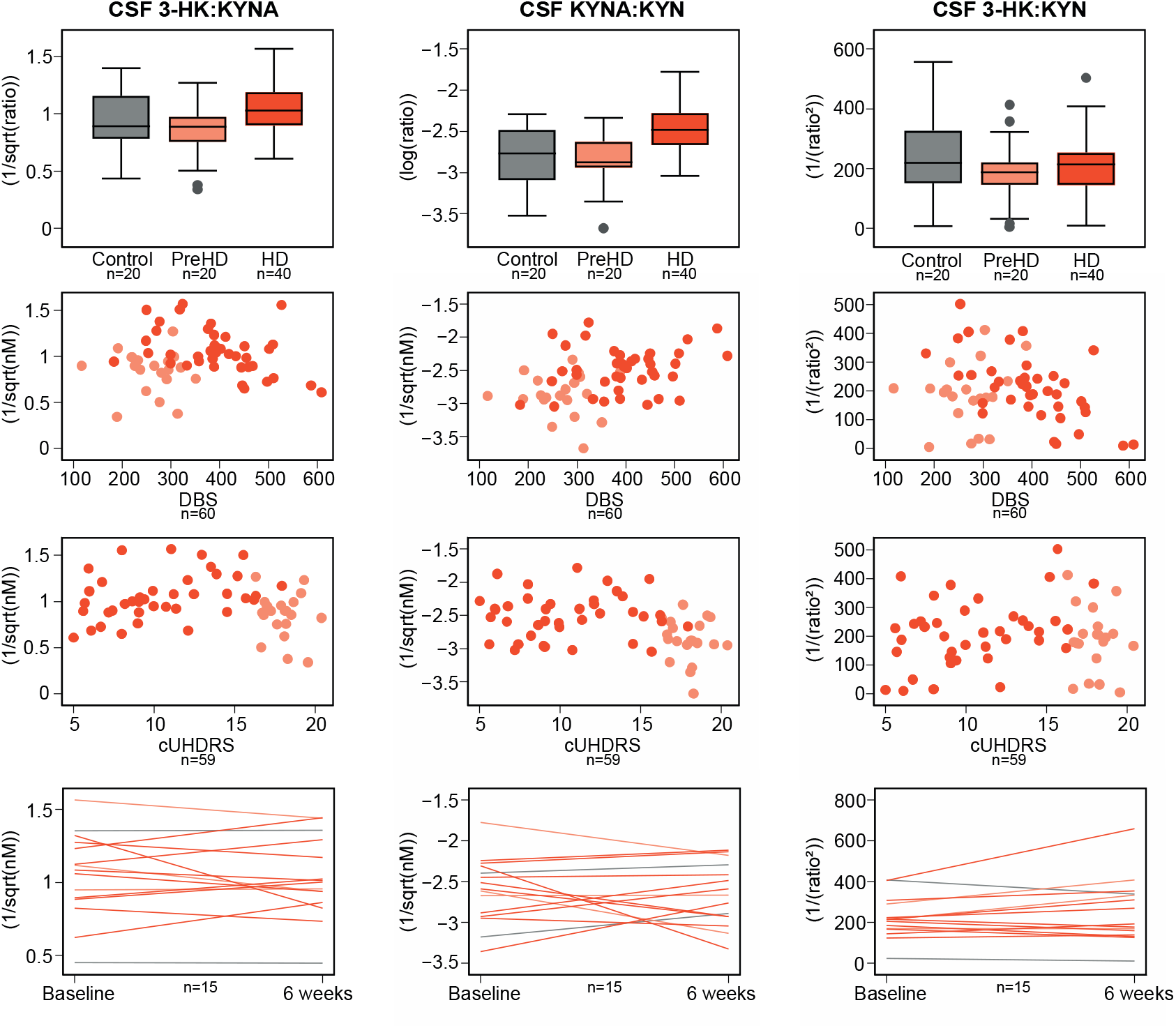
Intergroup differences (top row), associations with Disease Burden Score (DBS, second row) and composite Unified Huntington’s Disease Rating Scale (cUHDRS, third row), and within-subject short-term stability (bottom row) for secondary outcomes: ratios of cerebrospinal fluid (CSF) 3-hydroxykynurenine to kynurenic acid (3-HK:KYNA), CSF kynurenic acid to kynurenine (KYNA:KYN) and CSF 3-hydroxykynurenine to kynurenine (3-HK:KYN). Note that for outcomes where a transformation including a reciprocal (i.e. 1/x), an increase in the raw measurement corresponds to a decreased in the transformed value and vice-versa. Grey represents healthy controls, light orange preHD and dark orange HD.

Associations between age-adjusted CSF ratios and DBS, clinical and imaging measures were negligible to weak for 3-HK:KYNA, weak for KYNA:KYN, and weak to moderate for 3-HK:KYN (*Figure 4*, Table S2 and Figure S8).

Within-subject short-term stability was good for 3-HK:KYNA (ICC 0·78, 95% CI 0·46 to 0·92) and 3-HK:KYN (ICC 0·78, 95% CI 0·47 to 0·92), and weak to moderate for KYNA:KYN (ICC 0·43, 95% CI −0·09 to 0·76, *Figure 5* and Table S3).

Power calculations showed that the number of participants per arm needed to give 80% power to detect a difference at the alpha level of 005 in age-adjusted CSF levels between healthy controls and preHD would be 777 for 3-HK:KYNA, 13,521 for KYNA:KYN and 417 for 3-HK:KYN. For the comparison between preHD and HD the number would be 207 for 3-HK:KYNA, 126 for KYNA:KYN and 3,993 for 3-HK:KYN.

### Exploratory outcomes

Associations between age, gender, CAG repeat count, CSF haemoglobin or time in the freezer are shown in Figure S6. Age- and age and CAG-adjusted levels of CSF and plasma exploratory outcomes showed no difference between groups (Figure S7 and *Table 2)*. While plasma KYN, 3-HK, AA and QUIN showed negligible to weak associations with DBS, clinical and imaging measures; CSF AA showed negligible to moderate associations, and CSF TRP, KYN, and plasma TRP and KYNA showed weak to moderate associations (Figure S7, Figure S8 and Table S2). Within-subject short-term stability varied from low (plasma KYNA) to high (plasma QUIN) and is summarised in Table S3.

### Association between CSF and plasma

The matched CSF and plasma analytes were associated, with the exception of KYNA *(Figure 6* and Table S4). The strongest association was seen for QUIN (r = −0·75, 95% CI −0·83 to – 0·65).

**Figure 6.**
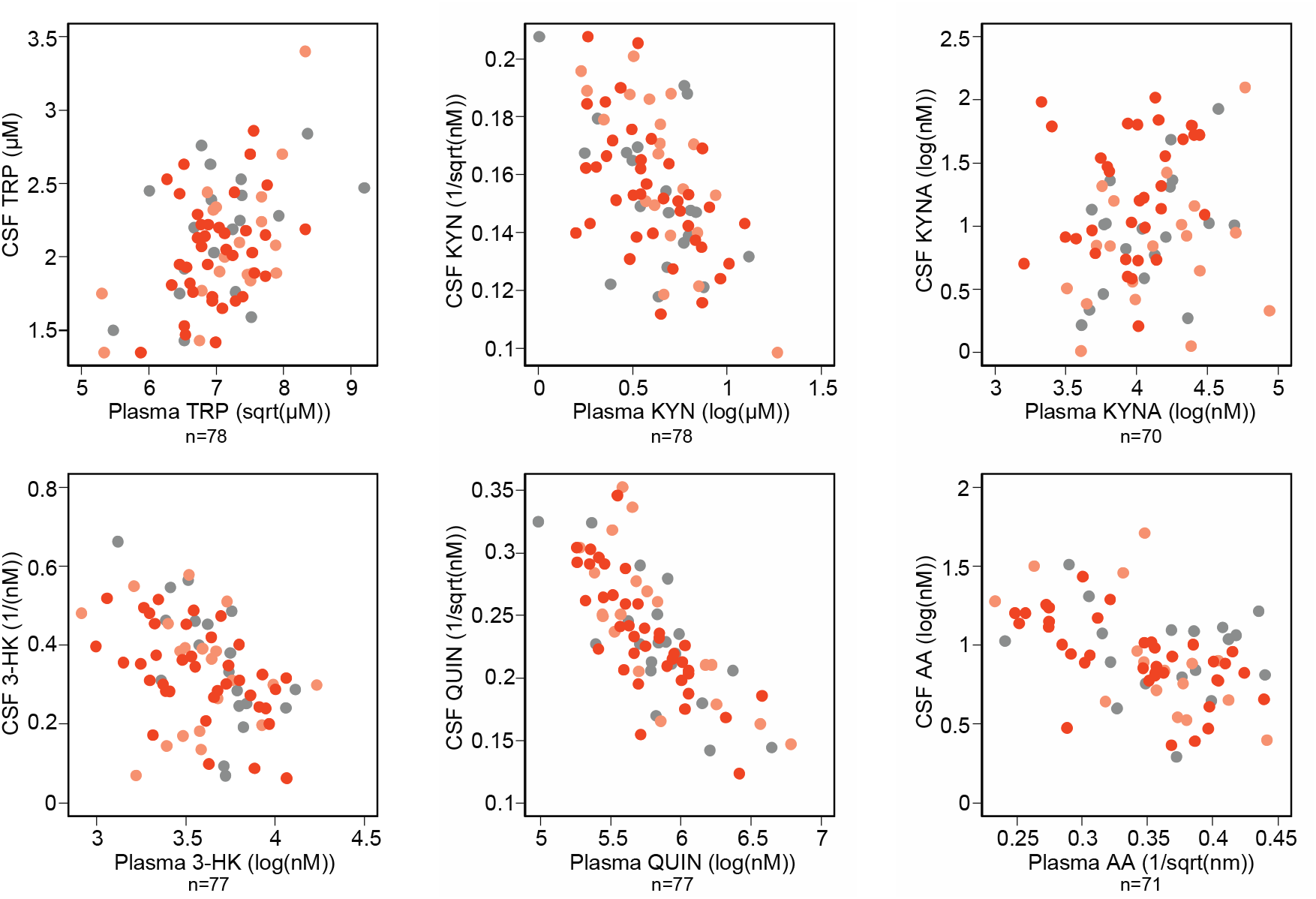
Associations between CSF and plasma. Grey represents healthy controls, light orange preHD and dark orange HD.

## Discussion

In this comprehensive study of KP metabolites in CSF and plasma in HD, we found that TRP, KYN, KYNA, 3-HK, AA and QUIN were readily and reliably quantifiable in both biofluids in controls and gene expansion carriers. However, we found little evidence to support a substantial derangement of the KP in HD, at least to the extent that it is reflected by the levels of the metabolites in patient-derived biofluids.

None of the three CSF metabolites we pre-specified as being of particular interest – KYNA, 3-HK and QUIN – had a significantly altered level in preHD or HD. We found no association between CSF levels of these metabolites and any robust clinical, cognitive and MR brain volumetric measures.

We did find significant associations between all three and age, which might (along with lesser assay reliability and sample quality) be one reason why previous studies reported some alterations, albeit inconsistently. Careful examination and adjustment for confounding variables is especially important given that HD is characterised by an extended premanifest period followed by a long period of manifest disease. This tends to make premanifest cohorts younger than those with manifest HD, and makes it very challenging to recruit a control group that matches the HD groups well for age. We previously showed that an apparent increase in CSF TREM2 in HD was in fact an artefact of its relationship with age (51).

We pre-defined three CSF metabolite ratios that might change in the presence of specific predicted derangements of the KP – 3-HK:KYNA as a reflection of the ratio of the main protective and neurotoxic metabolites whose synthetic enzymes have been implicated in HD; KYNA:KYN as a measure of the activity of the KATs and 3-HK:KYN as a measure of the activity of KMO. We found no evidence for altered CSF 3-HK:KYN ratio. While there was no evidence of CSF 3-HK:KYNA and KYNA:KYN ratio differences between healthy controls and pre-HD, HD seems to have lower 3-HK:KYNA ratio than preHD (shown as a higher 1/Vratio) and higher KYNA:KYN (shown as a higher log(ratio)), differences that remained significant after adjustment for age and CAG. Note that for outcomes where a transformation including a reciprocal (i.e. 1/x), an increase in the raw measurement corresponds to a decreased in the transformed value and vice-versa. This suggests that the activity of KATs may be increased in HD, an increase that appears to grow as the disease progresses. This may reflect a primary effect of the HD mutation on KATs, or compensatory overactivity because of alterations in the central KP.

As expected, the CSF and plasma levels of each metabolite were significantly associated, and the plasma levels of each were higher, in line with previous reports in Alzheimer’s disease and depression.(52, 53)

Though largely negative, our findings do not imply that the KP in general, or KMO in particular, is not a valid therapeutic target. It is possible that cell-specific or region-specific disease-related alterations in this pathway contribute substantially to the pathogenesis of HD and that pharmacologically correcting them could favourably modify the course of the disease. In that context, it is therefore still possible that measuring KP metabolite levels in CSF could provide one or more valuable readouts of target engagement or meaningful biological effect, for instance by increasing the level of protective substances to above the baseline or control level. The non-significant, small differences between groups could also prove robust if tested in a much larger sample set; we have offered sample size calculations for such an experiment. Overall, however, this study provides little support for a relevant alteration of kynurenine pathway function in HD that is biochemically detectable in accessible patient biofluids.

## Data Availability

The data that support the findings of this study are available on request from the corresponding author, EJW. The data are not publicly available due to their containing information that could compromise the privacy of research participants.

## Contributors

EJW designed the study with the input of FG. FBR and LMB were involved in participant recruitment. Eligibility, clinical examinations and sample collection were performed by FBR, LMB, and RT. Imaging assessments were conceived RIS, EBJ and EDV, data was acquired by EDV, MA, EBJ, and processed by RIS and EBJ. MM and GF processed and analysed the patient samples. FBR developed and performed the statistical analysis; FBR, AJL and EJW interpreted the data and wrote the manuscript; and all authors contributed to reviewing the manuscript.

## Acknowledgements

We would like to thank all the participants from the HD community who donated samples and gave their time to take part in this study. This study was supported by the Medical Research Council UK and CHDI foundation.

## Data Sharing Statement

Figures

Figure 1 Overview of the kynurenine pathway. Adapted with authors and editors’ permission (1). The designations ‘neurotoxic’ and ‘neuroprotective’ are assigned on the basis of the balance of evidence and are acknowledged to be simplifications of complex properties 5

Figure 1 Overview of the kynurenine pathway. Adapted with authors and editors’ permission (1). The designations ‘neurotoxic’ and ‘neuroprotective’ are assigned on the basis of the balance of evidence and are acknowledged to be simplifications of complex properties 5

*Figure 2 Intergroup differences (top row), associations with Disease Burden Score (DBS, second row) and composite Unified Huntington’s Disease Rating Scale (cUHDRS, third row), and within-subject short-term stability (bottom row) for primary outcomes: cerebrospinal fluid (CSF) kynurenic acid (KYNA), CSF 3-hydroxykynurenine (3-HK) and CSF quinolinic acid (QUIN). Associations were not found in any of the analyses. Grey represents healthy controls, light orange preHD and dark orange HD*. 12

*Figure 3 Associations between outcomes and clinical and imaging measures for primary, secondary and exploratory outcomes in gene expansion carriers. Numbers are Pearson’s partial correlations coefficients adjusted for age (top row), and for age and CAG (bottom row; see Table S2 for 95% CI and Figure S8 for scatter plots). 3-HK, 3-hydroxykynurenine; AA, Anthranilic acid; CSF, cerebrospinal fluid; cUHDRS, composite Unified Huntington’s Disease Rating Scale; KYN, kynurenine; KYNA, kynurenic acid; QUIN, quinolinic acid; SCN, Stroop Color Naming’ SDMT, Symbol Digit Modalities Test; SWR, Stroop Word Reading; TFC, UHDRS Total Functional Capacity; TMS, UHDRS Total Motor Score; TRP, tryptophan; VFC, Verbal Fluency – Categorical*. 15

*Figure 4 Intergroup differences (top row), associations with Disease Burden Score (DBS, second row) and composite Unified Huntington’s Disease Rating Scale (cUHDRS, third row), and within-subject short-term stability (bottom row) for secondary outcomes: ratios of cerebrospinal fluid (CSF) 3-hydroxykynurenine to kynurenic acid (3-HK:KYNA), CSF kynurenic acid to kynurenine (KYNA:KYN) and CSF 3-hydroxykynurenine to kynurenine (3- HK:KYN). Note that for outcomes where a transformation including a reciprocal (i.e. 1/x), an increase in the raw measurement corresponds to a decreased in the transformed value and vice-versa. Grey represents healthy controls, light orange preHD and dark orange HD*. 16

*Figure 5 Associations between CSF and plasma. Grey represents healthy controls, light orange preHD and dark orange HD*. 18

Tables

*Table 1 Chromatographic separation parameters for each metabolite and assay performance. Note that 2 manifest HD participants are missing plasma. 3-HK, 3-hydroxykynurenine; AA, anthranilic acid; ACN; acetonitrile; C, cerebrospinal fluid; FA, formic acid; H_2_O, ultra-purified water; KYN, kynurenine; KYNA, kynurenic acid; P, plasma; QUIN, quinolinic acid; TFA, trifluoroacetic acid; TRP, tryptophan*.

*Table 2 Intergroup comparisons. Note that for outcomes where a transformation including a reciprocal (i.e. 1/x), an increase in the raw measurement corresponds to a decreased in the transformed value and vice-versa. 3-HK, 3-hydroxykynurenine; AA, Anthranilic acid; CSF, cerebrospinal fluid; HC, healthy controls; KYN, kynurenine; KYNA, kynurenic acid; M, manifest HD; N/A, not applicable; PM, premanifest HD; QUIN, quinolinic acid; SD, standard deviation; TRP, tryptophan*.

## Supplementary material

### Supplementary tables

**Table S1.**
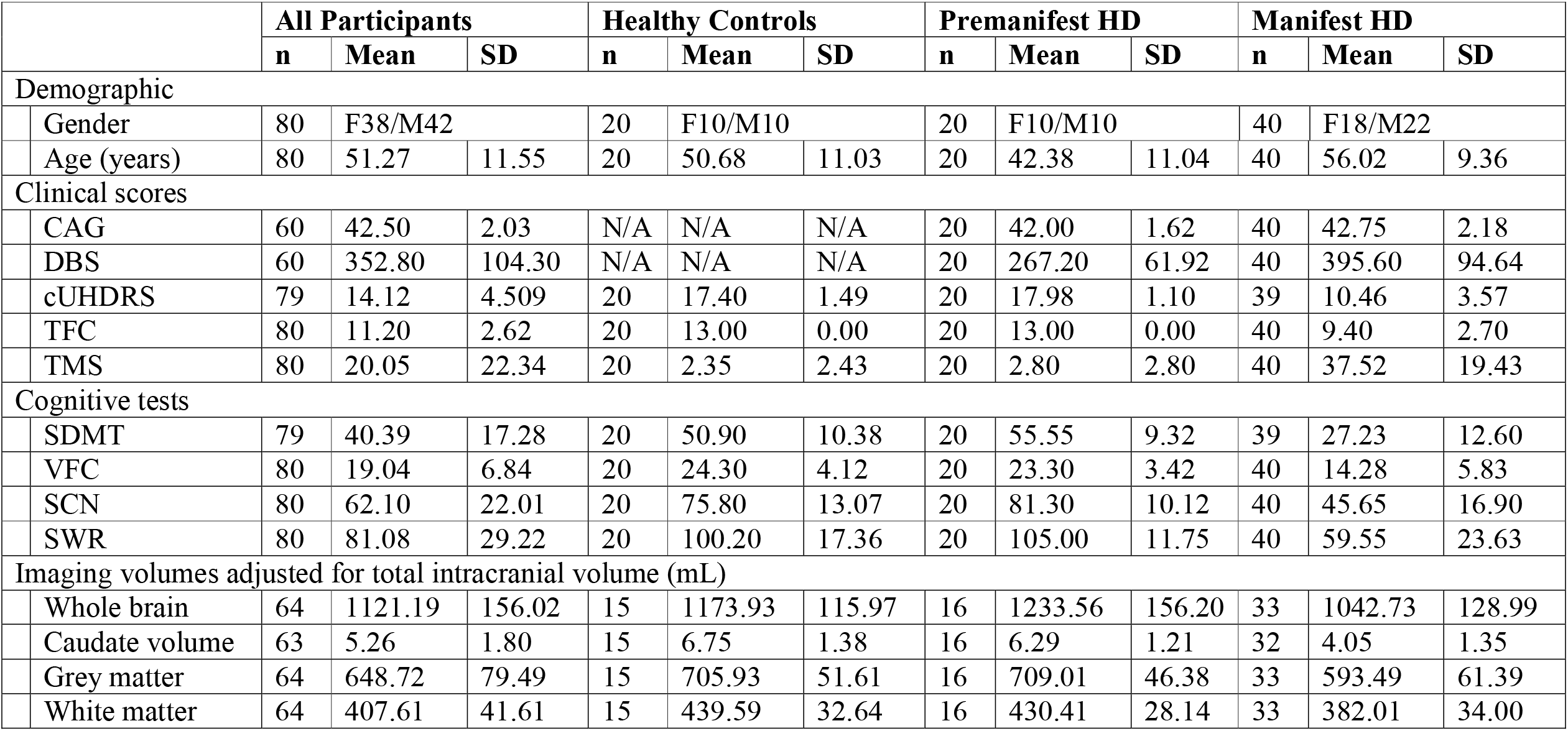
Cohort characteristics. CAG, CAG repeat count; DBS, Disease Burden Score; cUHDRS, composite Unified Huntington’s Disease Rating Scale; N/A, not applicable; SD, standard deviation; SDMT, Symbol Digit Modalities Test; SCN, Stroop Color Naming; SWR, Stroop Word Reading; TFC, UHDRS Total Functional Capacity; TMS, UHDRS Total Motor Score; VFC, Verbal Fluency – Categorical.

|  | All Participants |  |  | Healthy Controls |  |  | Premanifest HD |  |  | Manifest HD |  |  |
| --- | --- | --- | --- | --- | --- | --- | --- | --- | --- | --- | --- | --- |
|  | n | Mean | SD | n | Mean | SD | n | Mean | SD | n | Mean | SD |
| Demographic |  |  |  |  |  |  |  |  |  |  |  |  |
| Gender | 80 | F38/M42 |  | 20 | F10/M10 |  | 20 | F10/M10 |  | 40 | F18/M22 |  |
| Age (years) | 80 | 51.27 | 11.55 | 20 | 50.68 | 11.03 | 20 | 42.38 | 11.04 | 40 | 56.02 | 9.36 |
| Clinical scores |  |  |  |  |  |  |  |  |  |  |  |  |
| CAG | 60 | 42.50 | 2.03 | N/A | N/A | N/A | 20 | 42.00 | 1.62 | 40 | 42.75 | 2.18 |
| DBS | 60 | 352.80 | 104.30 | N/A | N/A | N/A | 20 | 267.20 | 61.92 | 40 | 395.60 | 94.64 |
| cUHDRS | 79 | 14.12 | 4.509 | 20 | 17.40 | 1.49 | 20 | 17.98 | 1.10 | 39 | 10.46 | 3.57 |
| TFC | 80 | 11.20 | 2.62 | 20 | 13.00 | 0.00 | 20 | 13.00 | 0.00 | 40 | 9.40 | 2.70 |
| TMS | 80 | 20.05 | 22.34 | 20 | 2.35 | 2.43 | 20 | 2.80 | 2.80 | 40 | 37.52 | 19.43 |
| Cognitive tests |  |  |  |  |  |  |  |  |  |  |  |  |
| SDMT | 79 | 40.39 | 17.28 | 20 | 50.90 | 10.38 | 20 | 55.55 | 9.32 | 39 | 27.23 | 12.60 |
| VFC | 80 | 19.04 | 6.84 | 20 | 24.30 | 4.12 | 20 | 23.30 | 3.42 | 40 | 14.28 | 5.83 |
| SCN | 80 | 62.10 | 22.01 | 20 | 75.80 | 13.07 | 20 | 81.30 | 10.12 | 40 | 45.65 | 16.90 |
| SWR | 80 | 81.08 | 29.22 | 20 | 100.20 | 17.36 | 20 | 105.00 | 11.75 | 40 | 59.55 | 23.63 |
| Imaging volumes adjusted for total intracranial volume (mL) |  |  |  |  |  |  |  |  |  |  |  |  |
| Whole brain | 64 | 1121.19 | 156.02 | 15 | 1173.93 | 115.97 | 16 | 1233.56 | 156.20 | 33 | 1042.73 | 128.99 |
| Caudate volume | 63 | 5.26 | 1.80 | 15 | 6.75 | 1.38 | 16 | 6.29 | 1.21 | 32 | 4.05 | 1.35 |
| Grey matter | 64 | 648.72 | 79.49 | 15 | 705.93 | 51.61 | 16 | 709.01 | 46.38 | 33 | 593.49 | 61.39 |
| White matter | 64 | 407.61 | 41.61 | 15 | 439.59 | 32.64 | 16 | 430.41 | 28.14 | 33 | 382.01 | 34.00 |

**Table S2.**
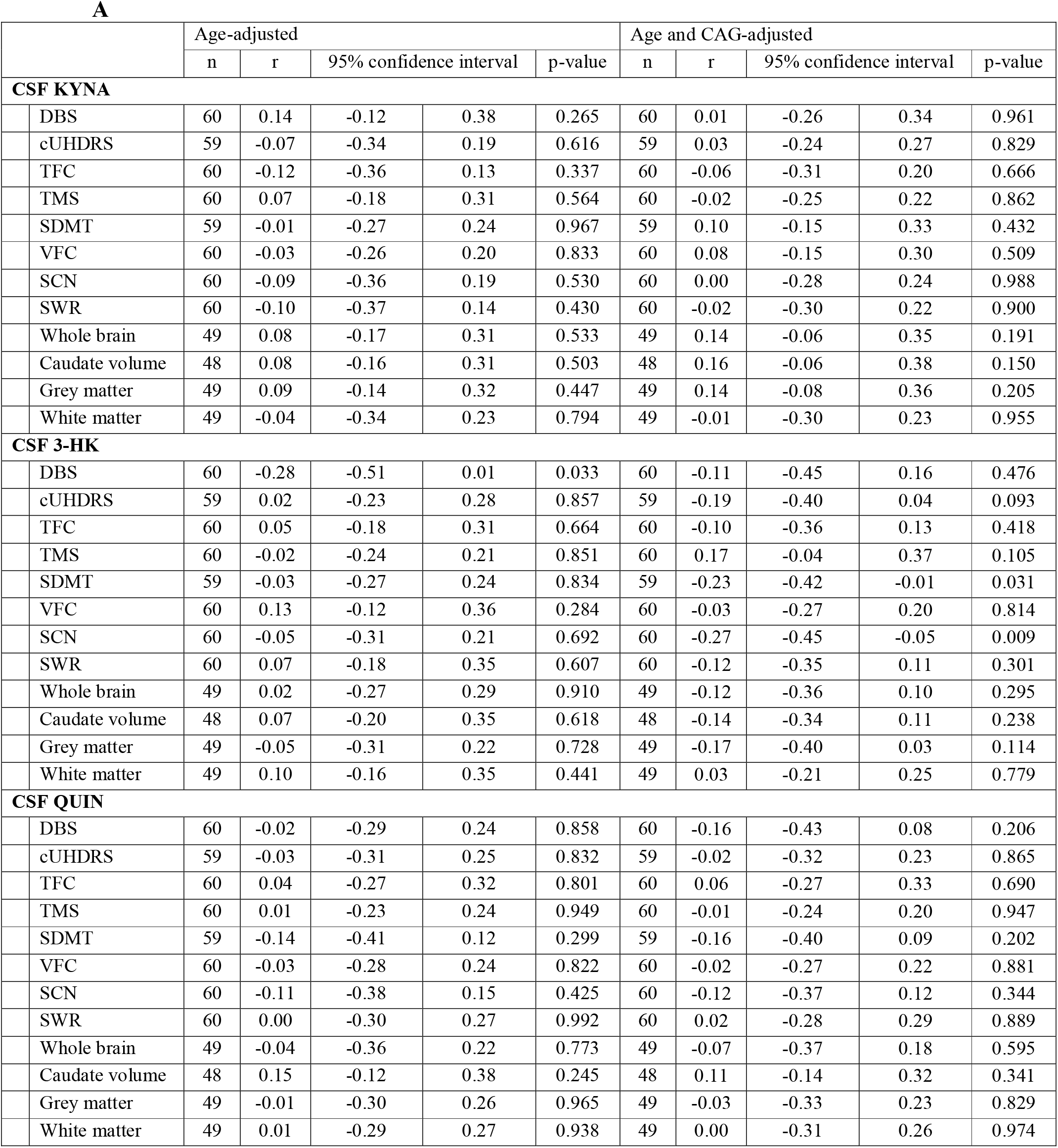
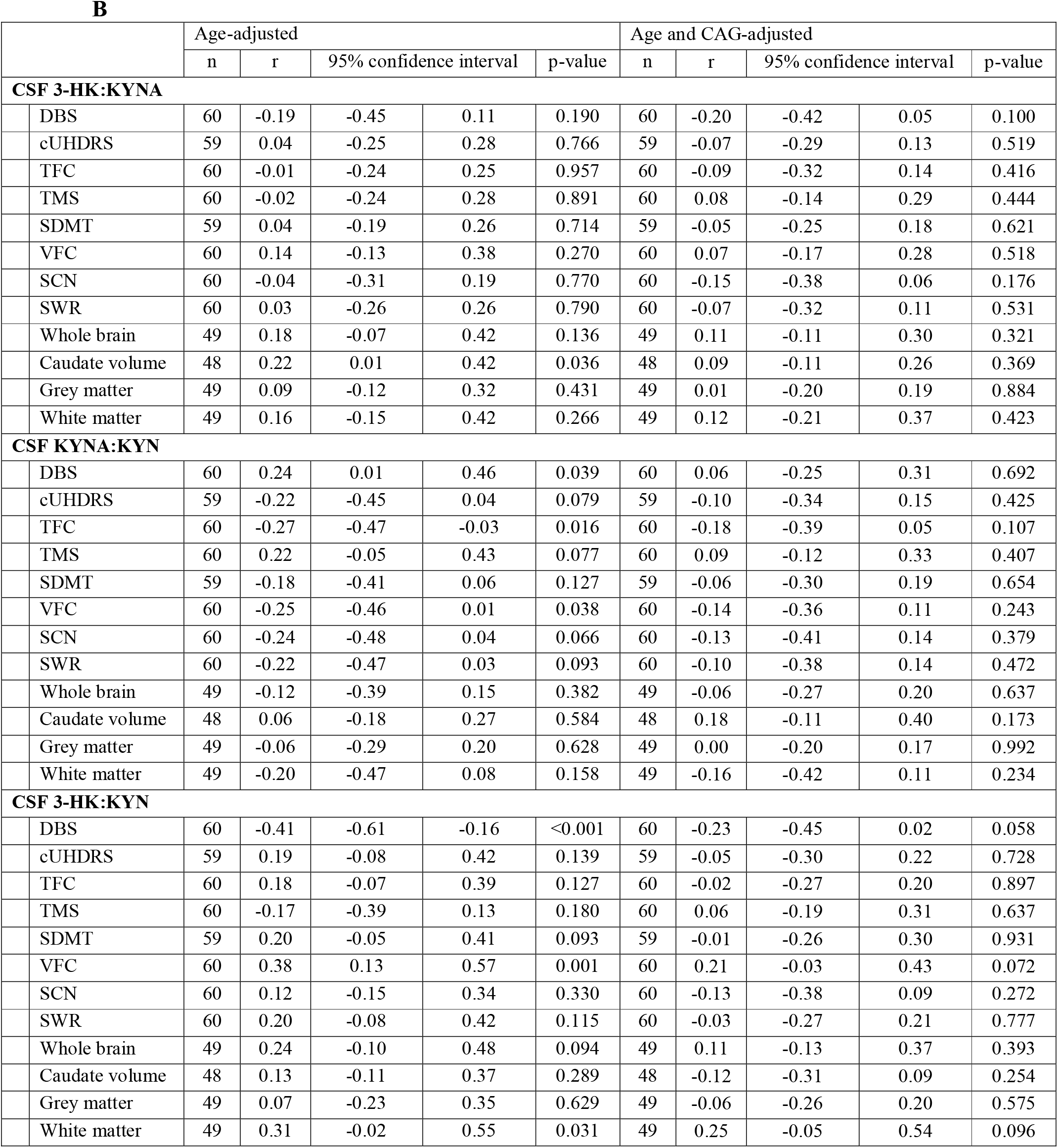
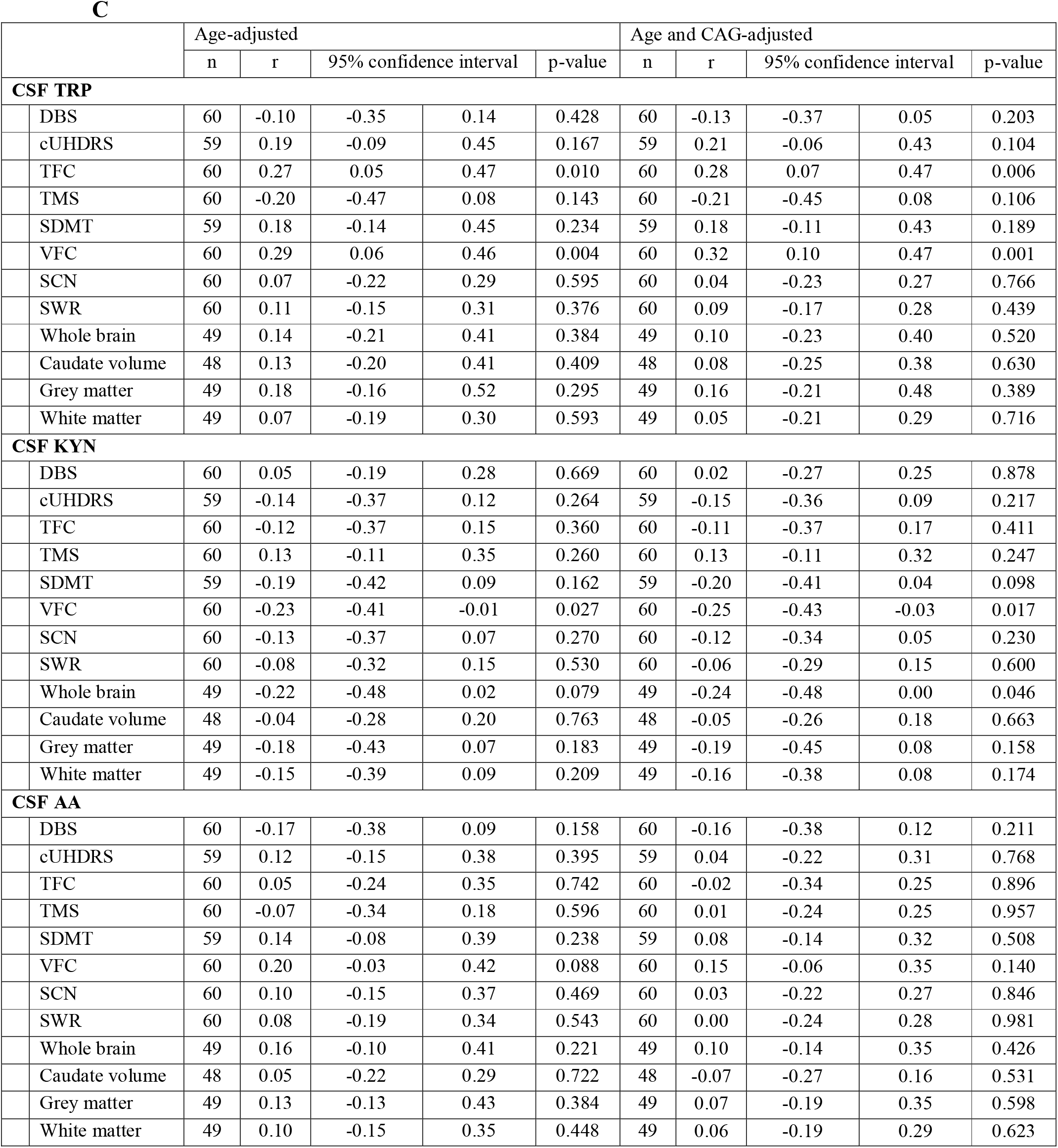
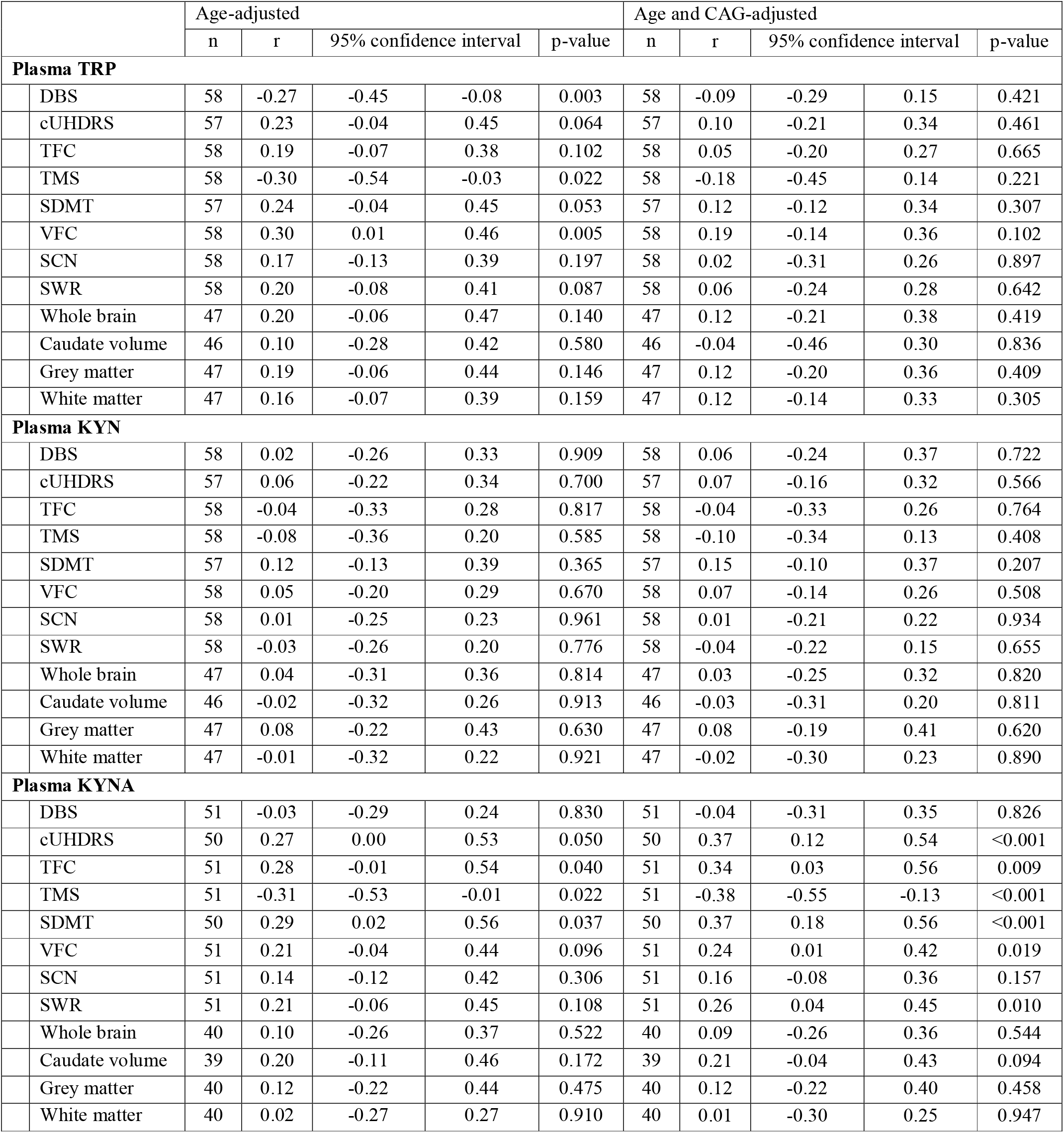
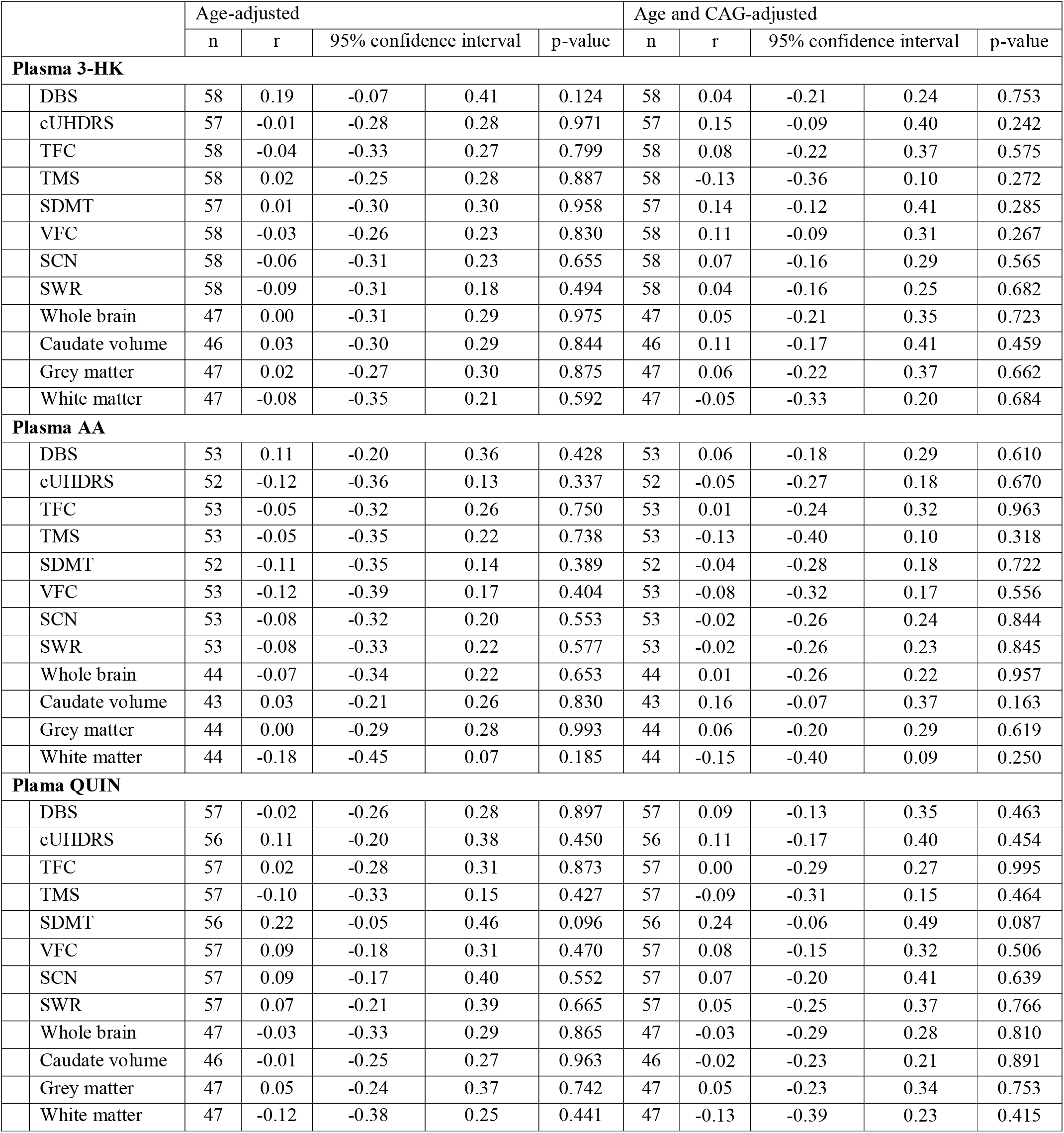
Associations between outcomes and disease burden score (DBS), clinical and imaging measures for (A) primary, (B) secondary and (C) exploratory outcomes in gene expansion carriers. 3-HK, 3-hydroxykynurenine; AA, Anthranilic acid; CSF, cerebrospinal fluid; cUHDRS, composite Unified Huntington’s Disease Rating Scale; KYN, kynurenine; KYNA, kynurenic acid; r, Pearson’s partial correlation coefficient; QUIN, quinolinic acid; SCN, Stroop Color Naming; SDMT, Symbol Digit Modalities Test; SWR, Stroop Word Reading; TFC, UHDRS Total Functional Capacity; TMS, UHDRS Total Motor Score; TRP, tryptophan; VFC, Verbal Fluency – Categorical.

**Table S3.** Within-subject short-term stability for primary, secondary and exploratory outcomes. 3-HK, 3-hydroxykynurenine; AA, Anthranilic acid; CSF, cerebrospinal fluid; ICC, KYN, kynurenine; KYNA, kynurenic acid; QUIN, quinolinic acid; TRP, tryptophan.

|  |  | ICC | 95% confidence interval |  |
| --- | --- | --- | --- | --- |
| Primary Outcomes |  |  |  |  |
|  | CSF KYNA | 0.84 | 0.58 | 0.94 |
|  | CSF 3-HK | 0.68 | 0.27 | 0.88 |
|  | CSF QUIN | 0.90 | 0.72 | 0.96 |
| Secondary Outcomes |  |  |  |  |
|  | CSF 3-HK:KYNA | 0.78 | 0.46 | 0.92 |
|  | CSF KYNA:KYN | 0.43 | -0.09 | 0.76 |
|  | CSF 3-HK:KYN | 0.78 | 0.47 | 0.92 |
| Exploratory Outcomes |  |  |  |  |
|  | CSF TRP | 0.58 | 0.11 | 0.83 |
|  | CSF KYN | 0.67 | 0.25 | 0.87 |
|  | CSF AA | 0.84 | 0.58 | 0.94 |
|  | Plasma TRP | 0.58 | 0.09 | 0.84 |
|  | Plasma KYN | 0.54 | 0.03 | 0.82 |
|  | Plasma KYNA | -0.16 | -0.68 | 0.45 |
|  | Plasma 3-HK | 0.57 | 0.08 | 0.84 |
|  | Plasma AA | 0.80 | 0.41 | 0.94 |
|  | Plasma QUIN | 0.89 | 0.69 | 0.96 |

**Table S4.**
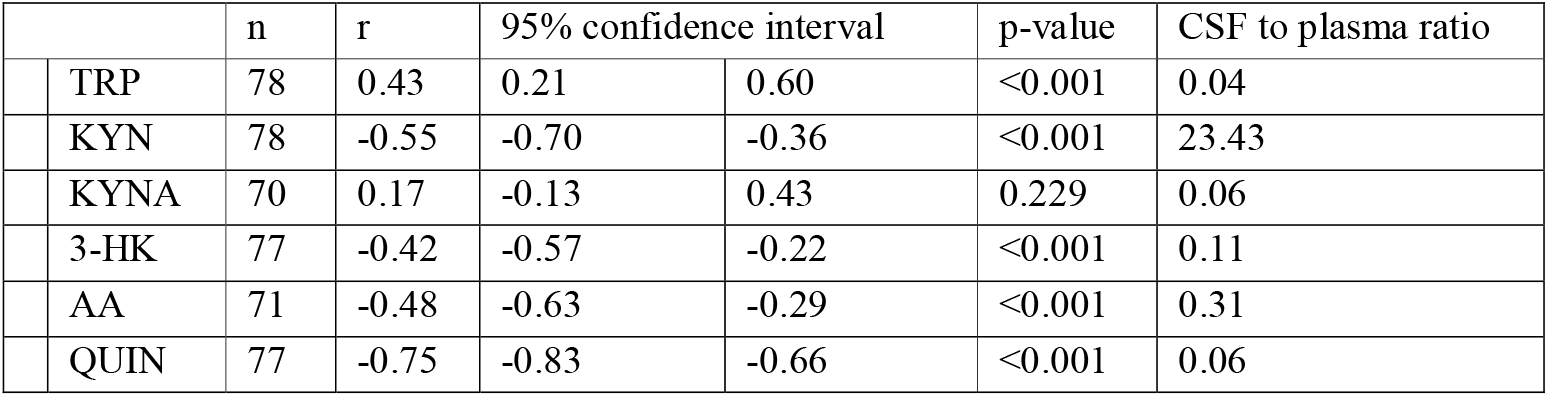
Associations and ratios between CSF and plasma. 3-HK, 3-hydroxykynurenine; AA, Anthranilic acid, KYN, kynurenine; KYNA, kynurenic acid; QUIN, quinolinic acid; r, Pearson’s partial correlation coefficient; TRP, tryptophan.

|  | n | r | 95% confidence interval |  | p-value | CSF to plasma ratio |
| --- | --- | --- | --- | --- | --- | --- |
| TRP | 78 | 0.43 | 0.21 | 0.60 | <0.001 | 0.04 |
| KYN | 78 | -0.55 | -0.70 | -0.36 | <0.001 | 23.43 |
| KYNA | 70 | 0.17 | -0.13 | 0.43 | 0.229 | 0.06 |
| 3-HK | 77 | -0.42 | -0.57 | -0.22 | <0.001 | 0.11 |
| AA | 71 | -0.48 | -0.63 | -0.29 | <0.001 | 0.31 |
| QUIN | 77 | -0.75 | -0.83 | -0.66 | <0.001 | 0.06 |

Supplementary figures

**Figure S1.**
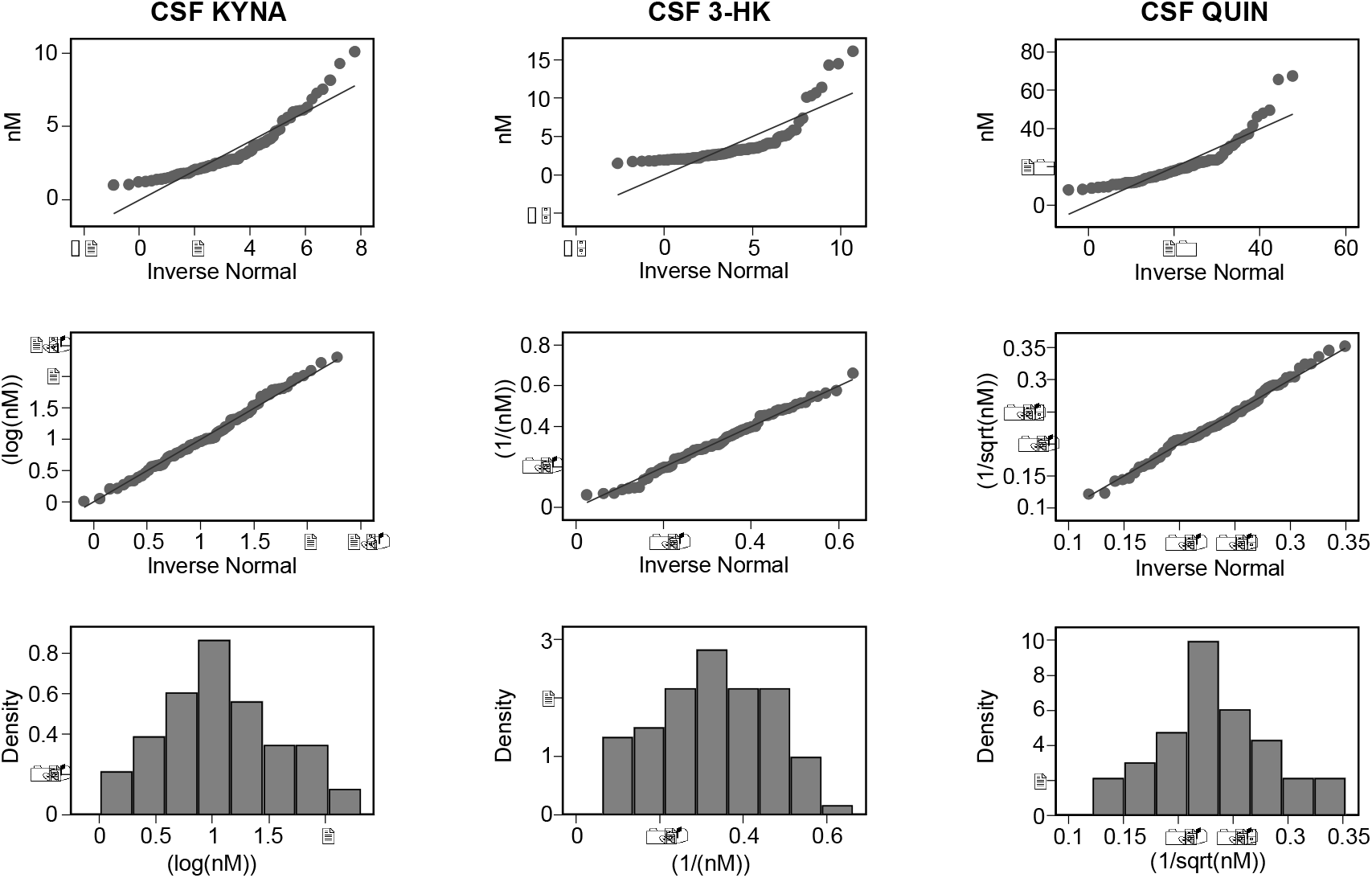
Distributions and arithmetic operations used to transform primary outcomes: cerebrospinal fluid (CSF) kynurenic acid (KYNA), CSF 3-hydroxykynurenine (3-HK) and CSF quinolinic acid (QUIN). Top row depicts a quantile-quantile plot for observed distribution against normal distribution, middle row shows a quantile-quantile plot for transformed distribution against normal distribution, and the bottom row a histogram for transformed distribution.

**Figure S2.**
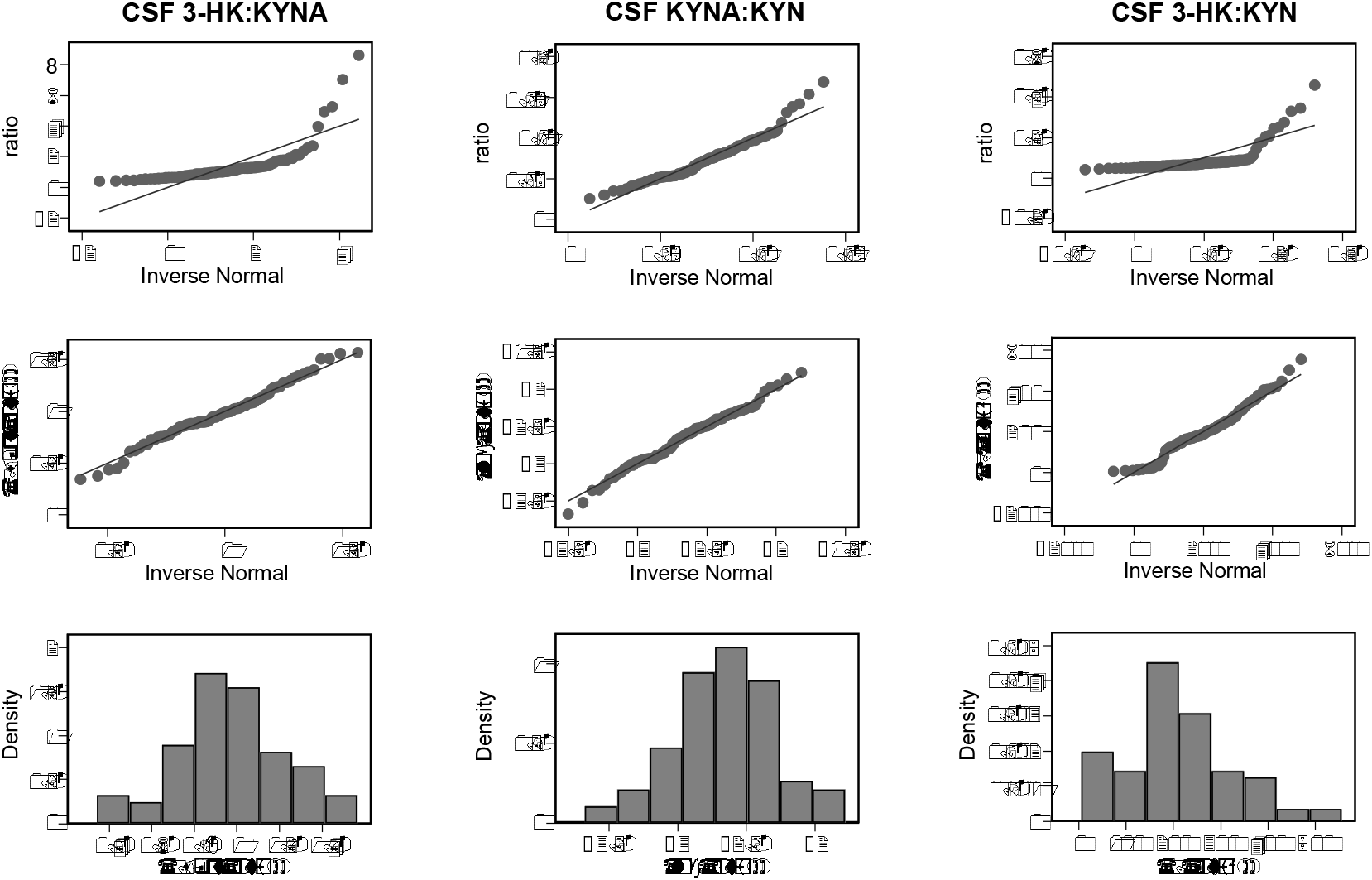
Distributions and arithmetic operations used to transform secondary outcomes: ratios of cerebrospinal fluid (CSF) 3-hydroxykynurenine to kynurenic acid (3-HK:KYNA), CSF kynurenic acid to kynurenine (KYNA:KYN) and CSF 3-hydroxykynurenine to kynurenine (3-HK:KYN). Top row depicts a quantile-quantile plot for observed distribution against normal distribution, middle row shows a quantile-quantile plot for transformed distribution against normal distribution, and the bottom row a histogram for transformed distribution.

**Figure S3.**
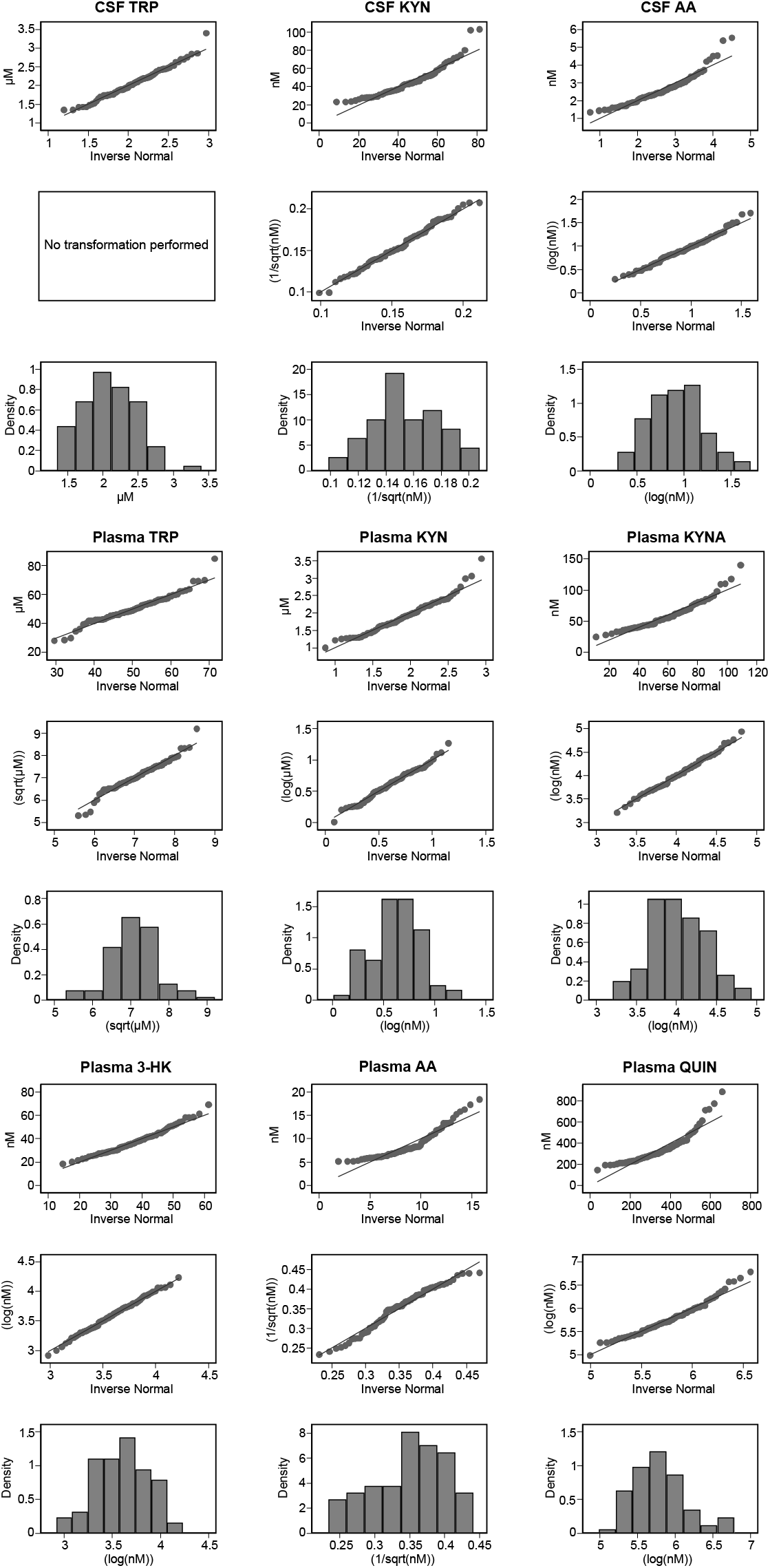
Distributions and arithmetic operations used to transform exploratory outcomes: cerebrospinal fluid (CSF) tryptophan (TRP), CSF kynurenine (KYN), CSF anthranilic acid (AA), plasma TRP, plasma KYN, plasma KYNA, plasma 3-hydroxykynurenine (3-HK), plasma AA, and plasma quinolinic acid (QUIN). In each of the three 3 by 3 subpanels, top row depicts a quantile-quantile plot for observed distribution against normal distribution, middle row shows a quantile-quantile plot for transformed distribution against normal distribution, and the bottom row a histogram for transformed distribution.

**Figure S4.**
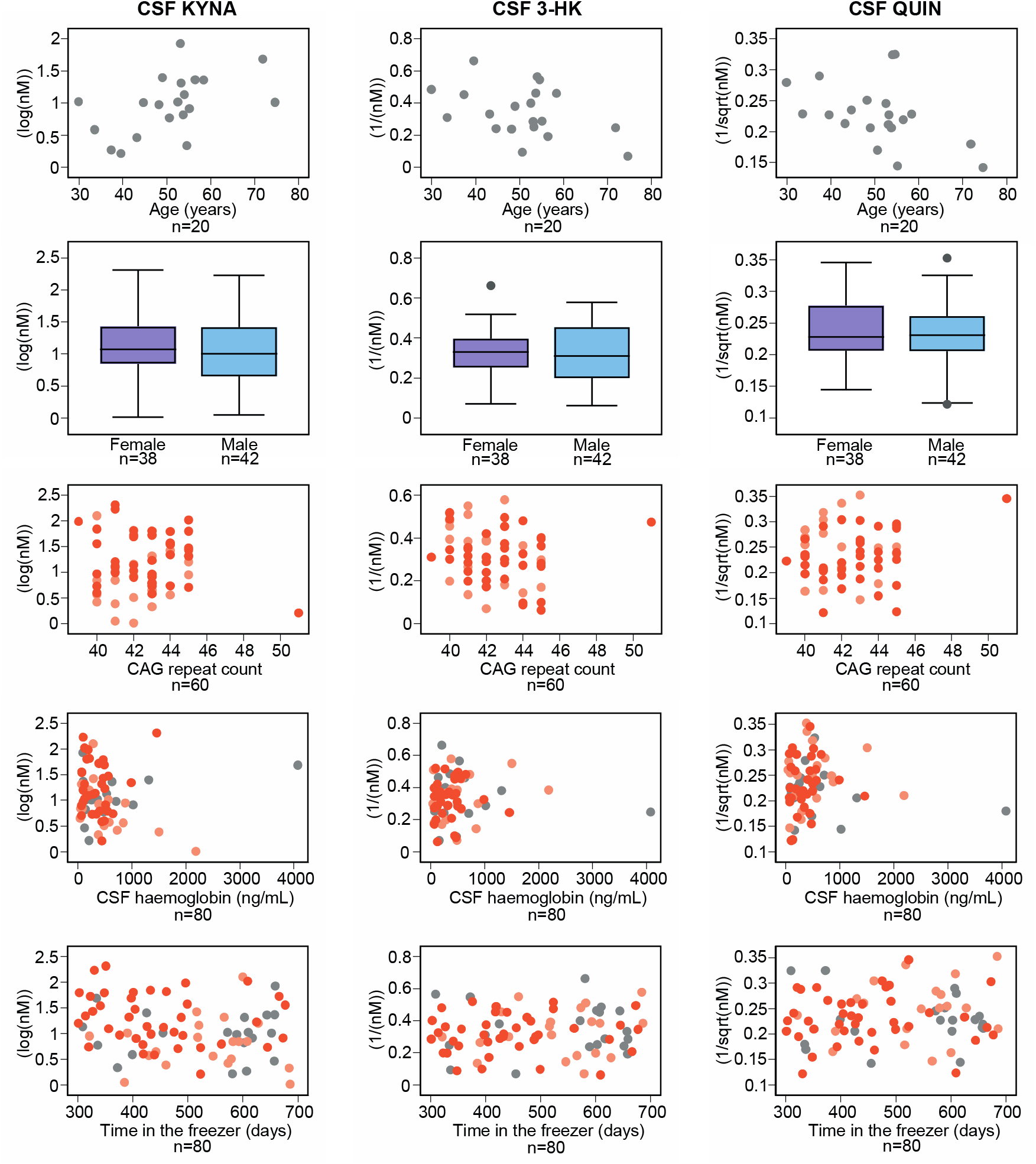
Associations between age (healthy controls only, top row), gender (second row), CAG repeat count (gene expansion carriers only, middle row), CSF haemoglobin (third row) and time in the freezer (bottom row) and primary outcomes: cerebrospinal fluid (CSF) kynurenic acid (KYNA), CSF 3-hydroxykynurenine (3-HK) and CSF quinolinic acid (QUIN). Grey represents healthy controls, light orange preHD and dark orange HD.

**Figure S5.**
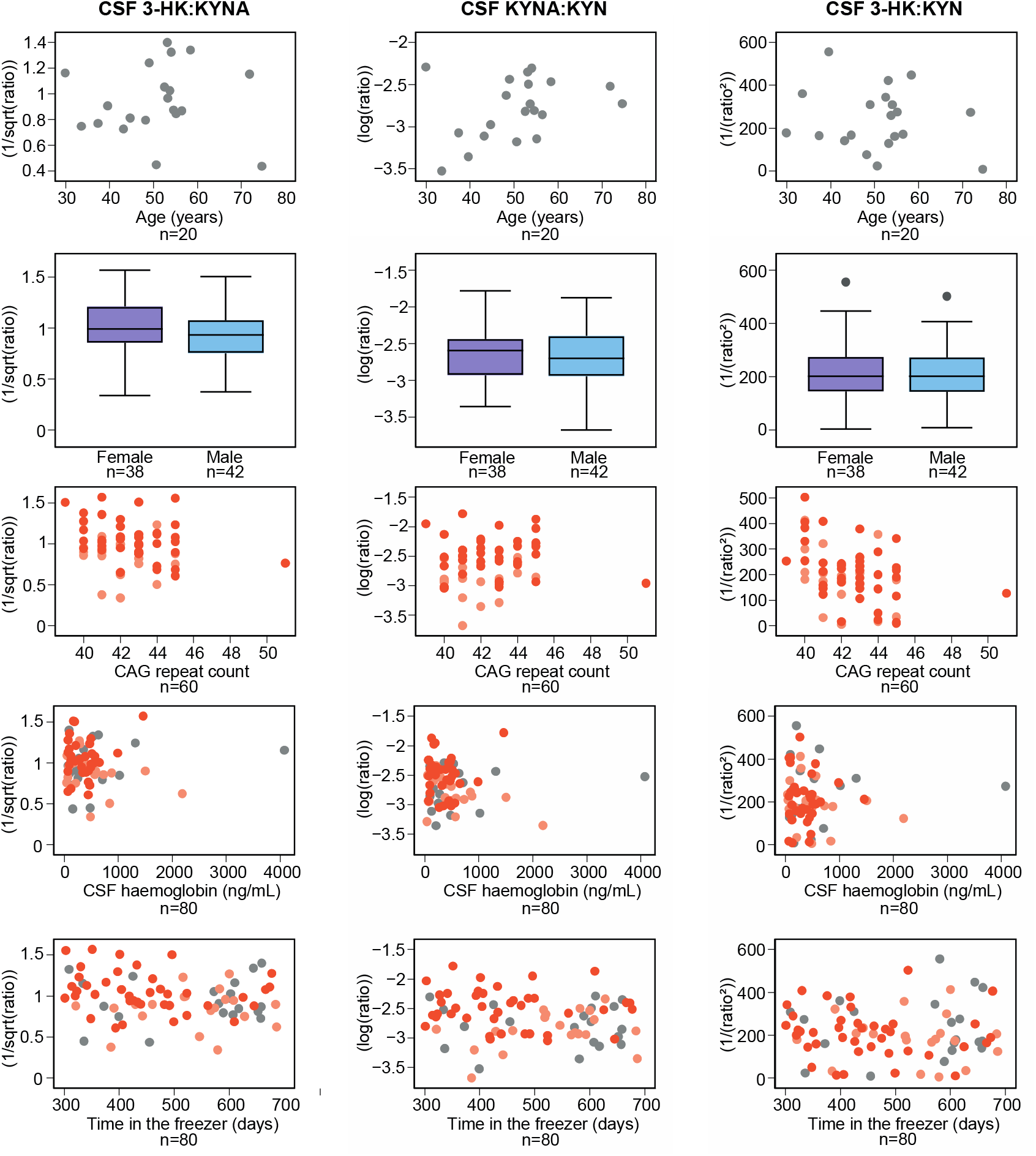
Associations between age (healthy controls only, top row), gender (second row), CAG repeat count (gene expansion carriers only, middle row), CSF haemoglobin (third row) and time in the freezer (bottom row) and secondary outcomes: ratios of cerebrospinal fluid (CSF) 3-hydroxykynurenine to kynurenic acid (3-HK:KYNA), CSF kynurenic acid to kynurenine (KYNA:KYN) and CSF 3-hydroxykynurenine to kynurenine (3-HK:KYN). Grey represents healthy controls, light orange preHD and dark orange HD.

**Figure S6.**
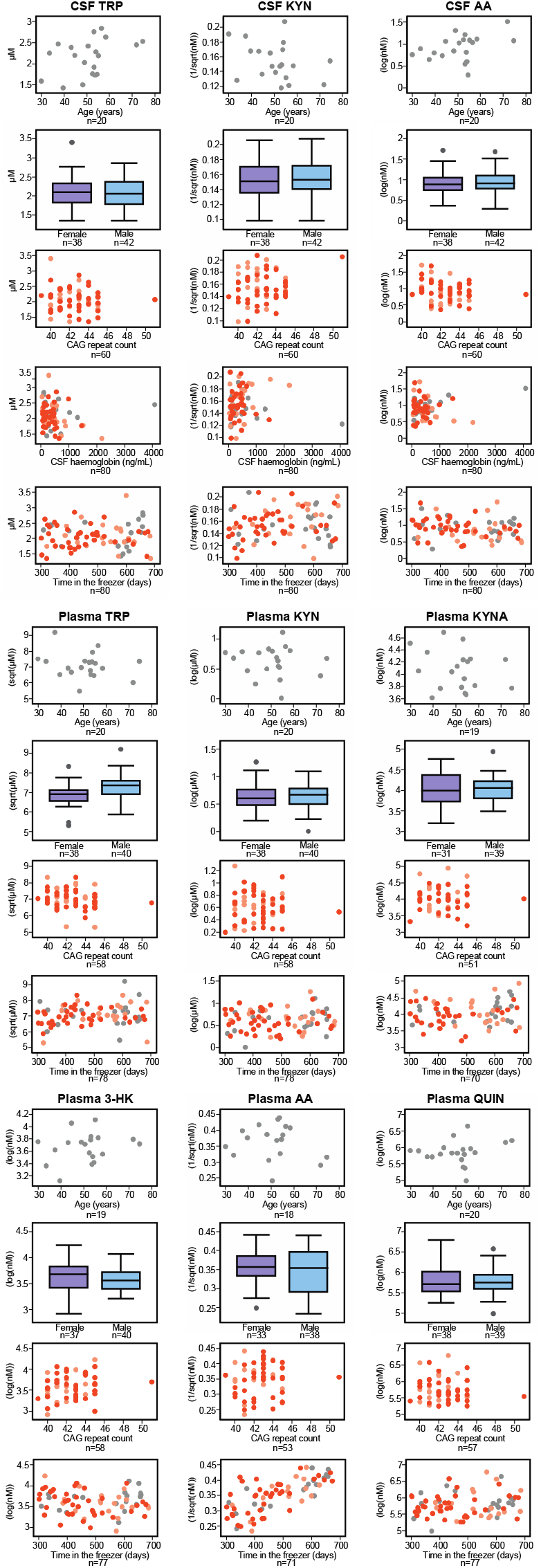
Associations between age (healthy controls only, top row), gender (second row), CAG repeat count (gene expansion carriers only, middle row), CSF haemoglobin (third row) and time in the freezer (bottom row) and exploratory outcomes: cerebrospinal fluid (CSF) tryptophan (TRP), CSF kynurenine (KYN), CSF anthranilic acid (AA), plasma TRP, plasma KYN, plasma KYNA, plasma 3-hydroxykynurenine (3-HK), plasma AA, and plasma quinolinic acid (QUIN). Grey represents healthy controls, light orange preHD and dark orange HD.

**Figure S7.**
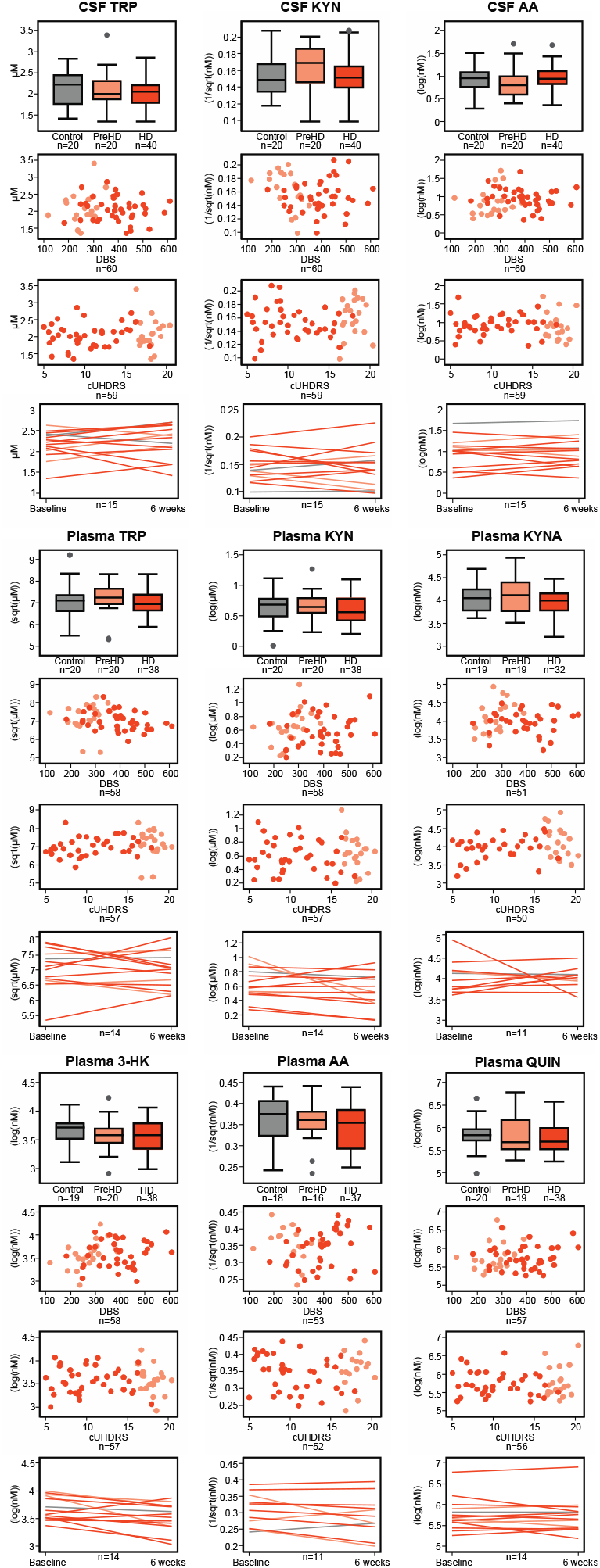
Intergroup differences (top row), associations with Disease Burden Score (DBS, second row) and composite Unified Huntington’s Disease Rating Scale (cUHDRS, third row), and within-subject short-term stability (bottom row) for exploratory outcomes: cerebrospinal fluid (CSF) tryptophan (TRP), CSF kynurenine (KYN), CSF anthranilic acid (AA), plasma TRP, plasma KYN, plasma KYNA, plasma 3-hydroxykynurenine (3-HK), plasma AA, and plasma quinolinic acid (QUIN). Grey represents healthy controls, light orange preHD and dark orange HD.

**Figure S8.**
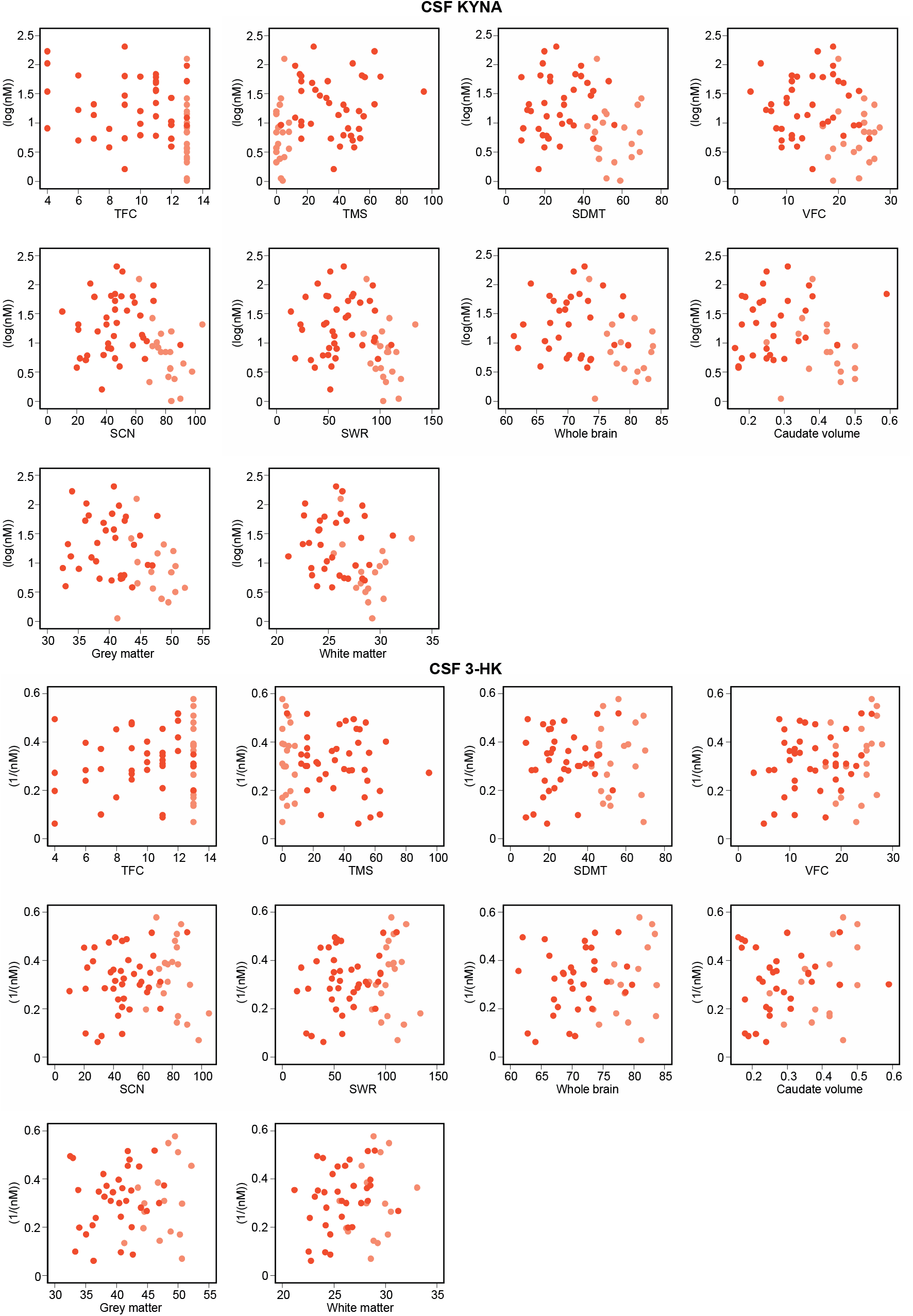
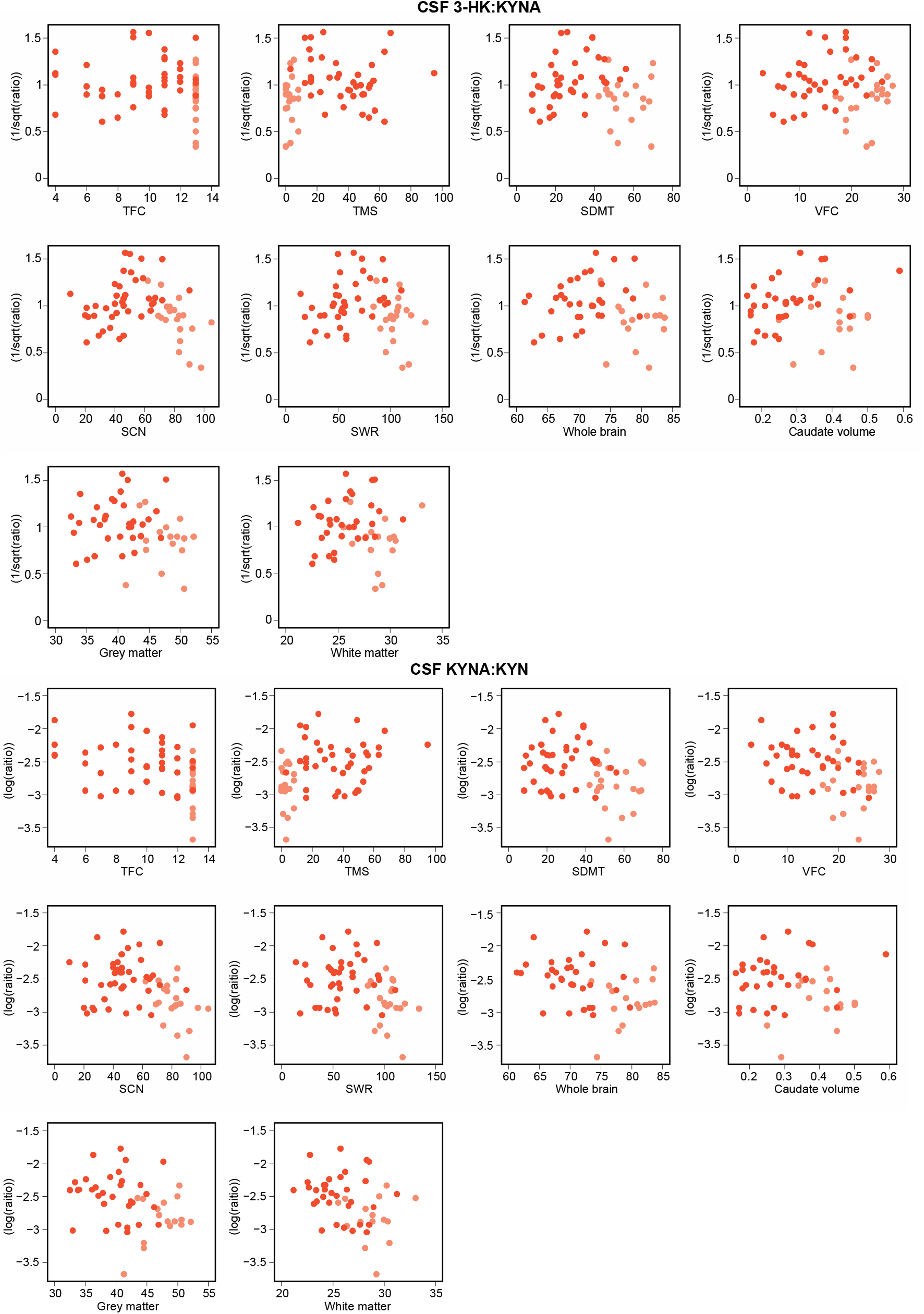
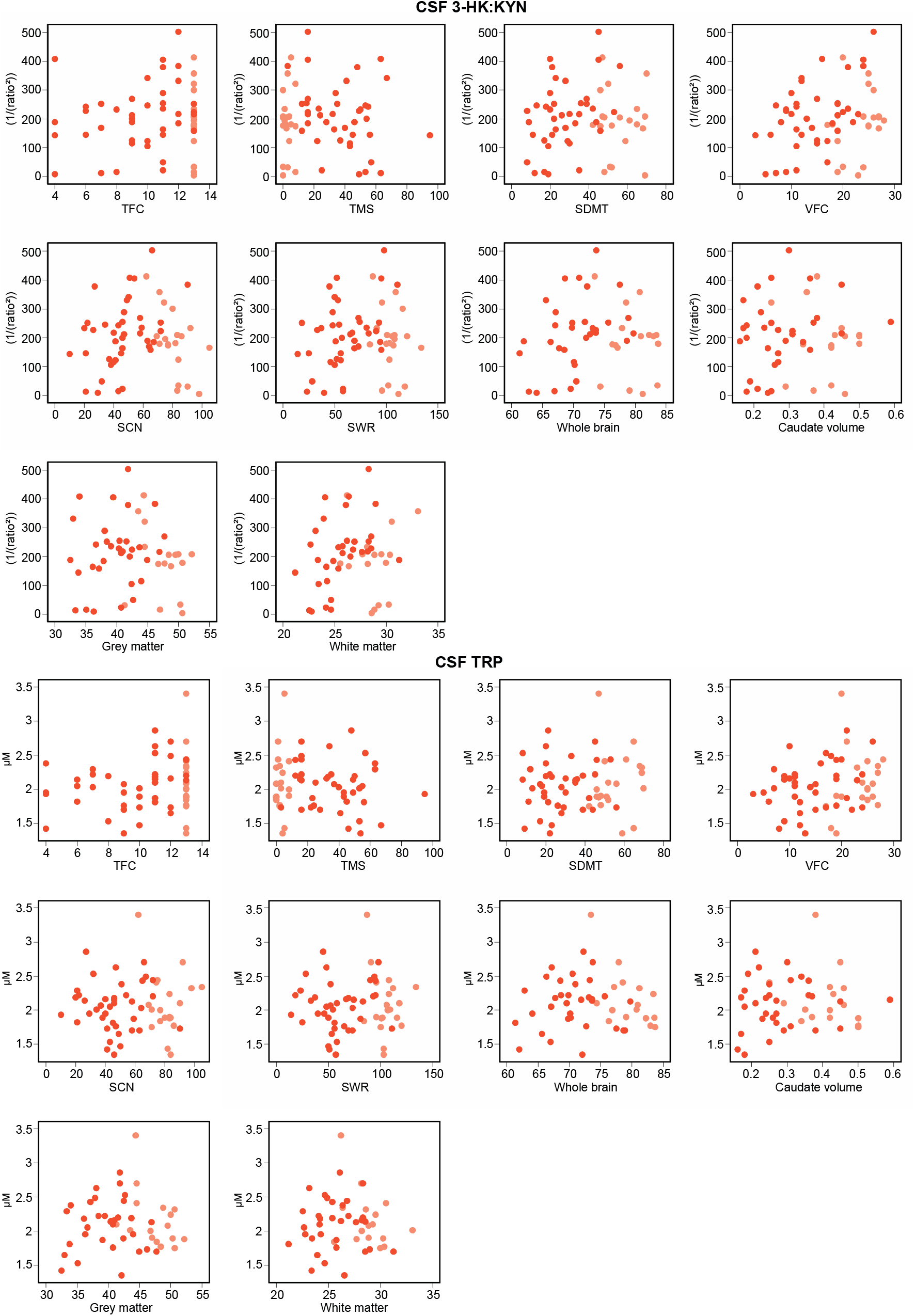
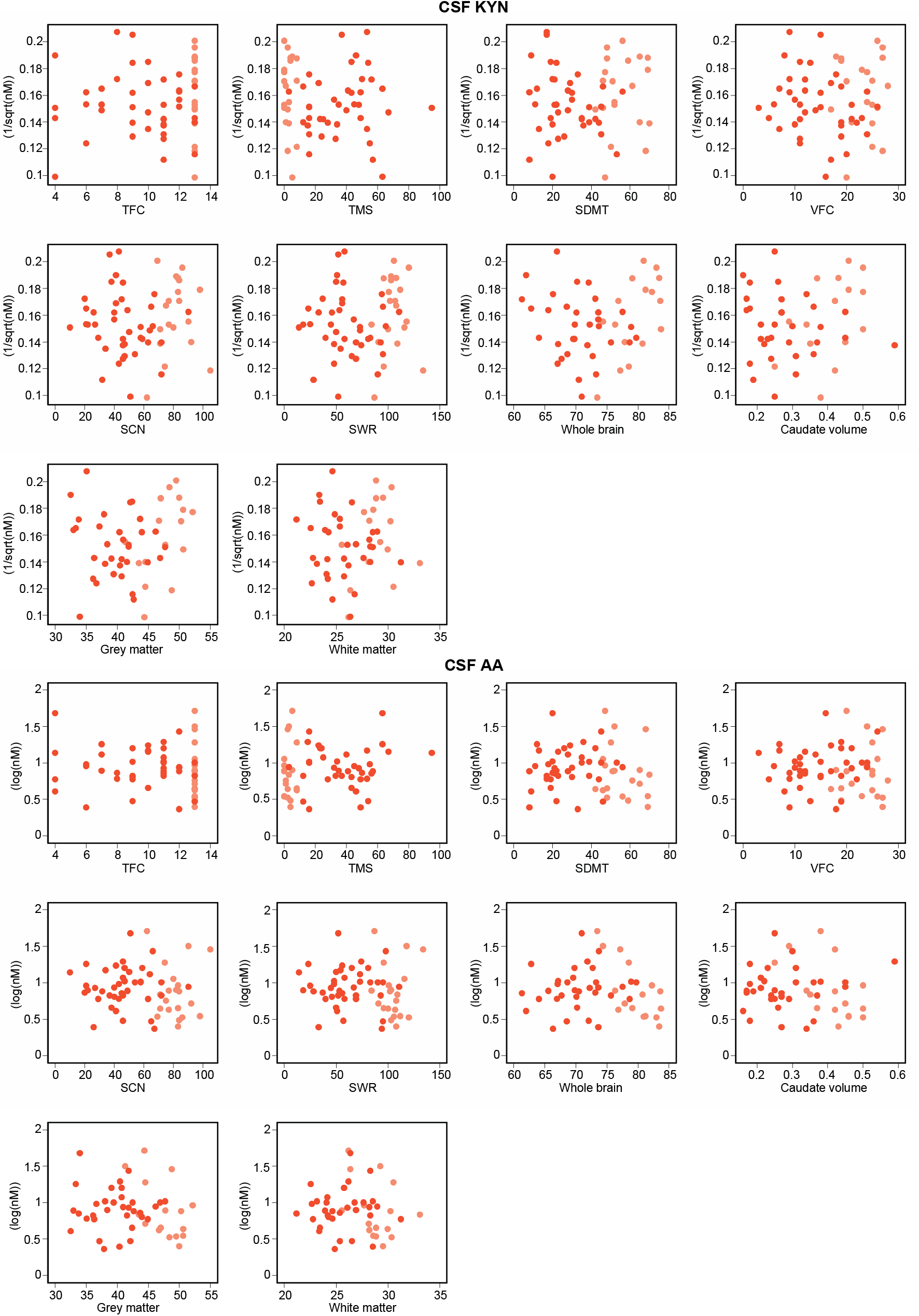
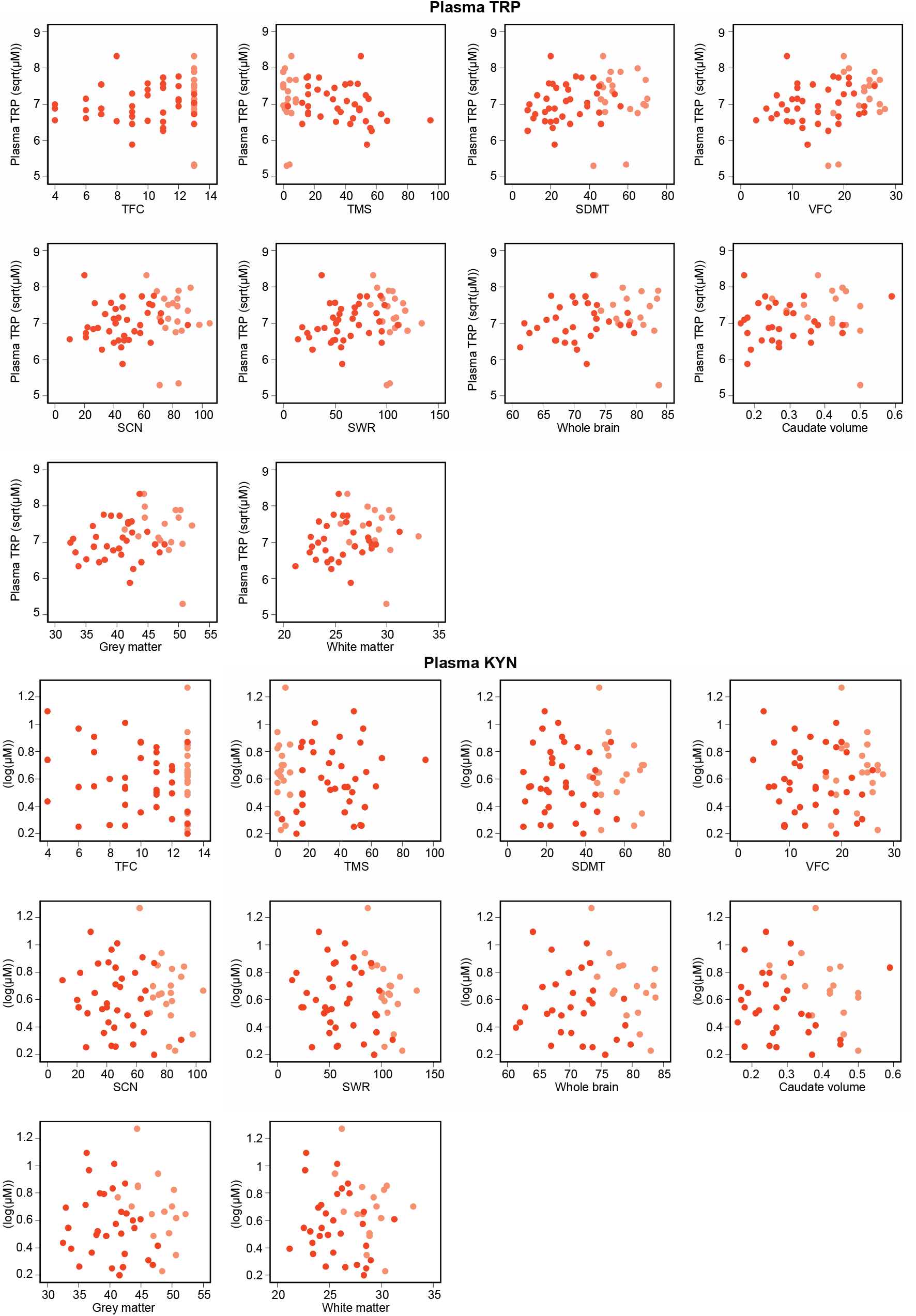
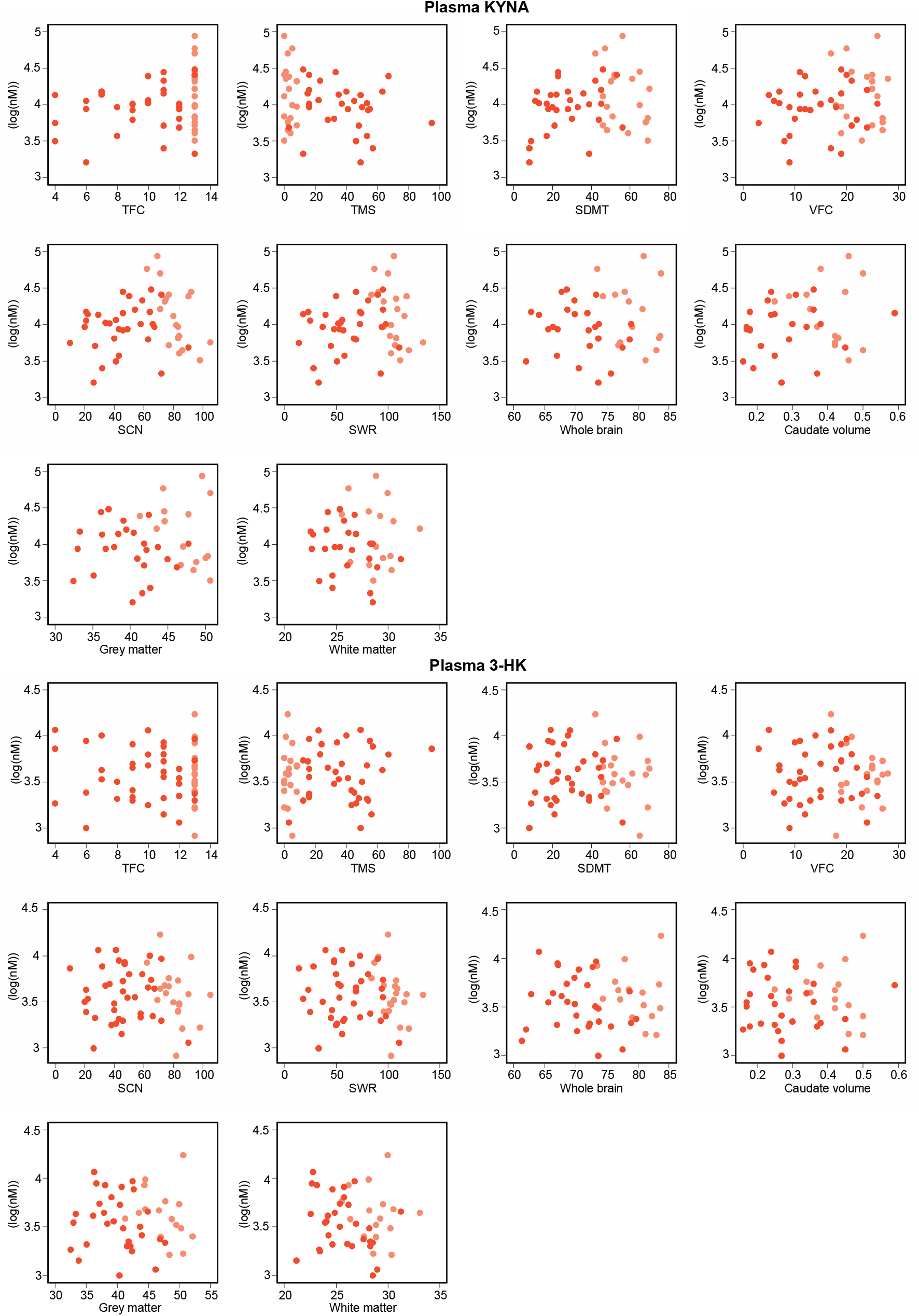
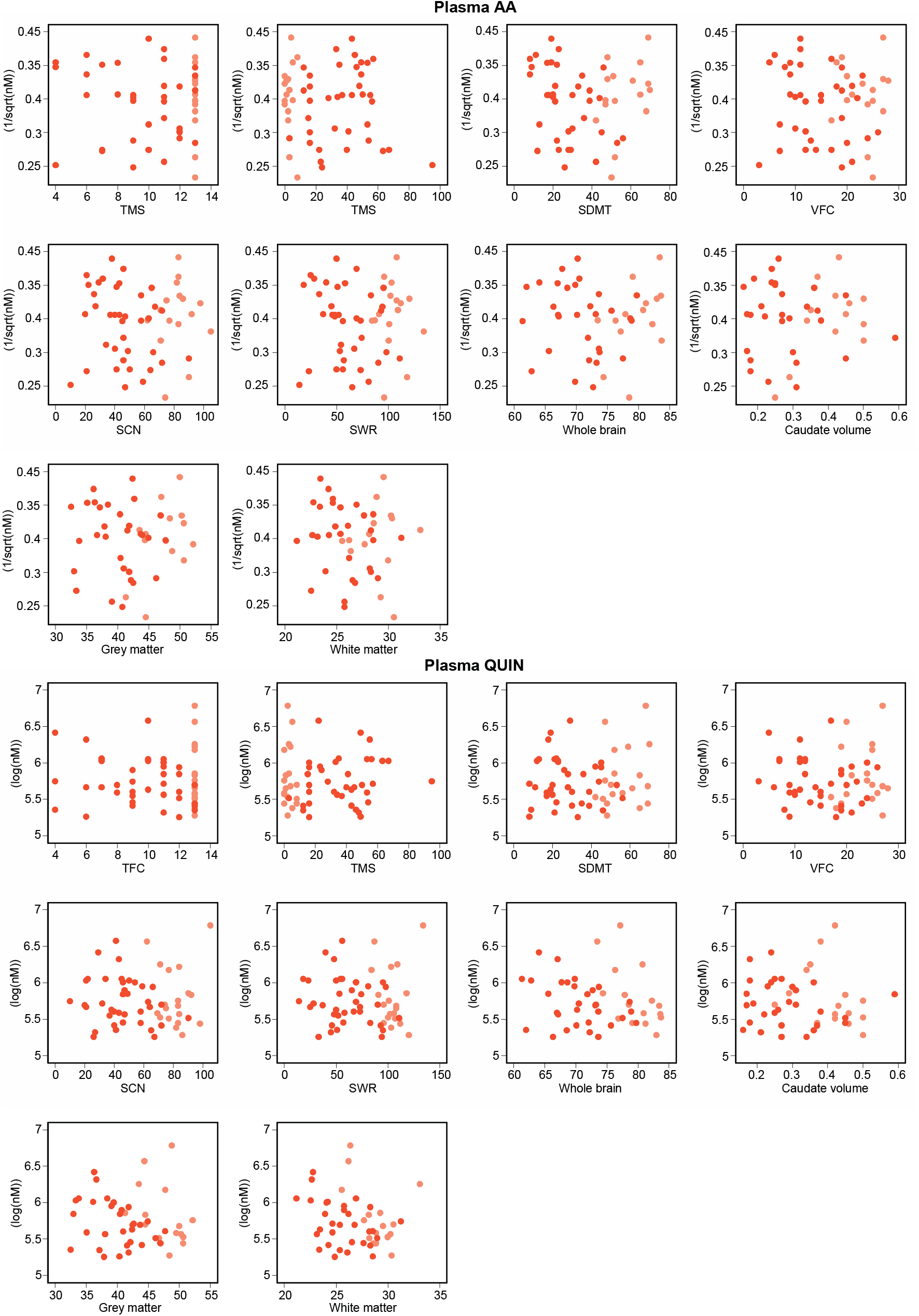
Associations between primary, secondary and exploratory outcomes and clinical and imaging measures. Grey represents healthy controls, light orange preHD and dark orange HD. 3-HK, 3-hydroxykynurenine; AA, Anthranilic acid; CSF, cerebrospinal fluid; cUHDRS, composite Unified Huntington’s Disease Rating Scale; KYN, kynurenine; KYNA, kynurenic acid; r, Pearson’s partial correlation coefficient; QUIN, quinolinic acid; SCN, Stroop Color Naming; SDMT, Symbol Digit Modalities Test; SWR, Stroop Word Reading; TFC, UHDRS Total Functional Capacity; TMS, UHDRS Total Motor Score; TRP, tryptophan; VFC, Verbal Fluency – Categorical.

